# Genetic discovery using deep learning-derived optic nerve integrity phenotypes

**DOI:** 10.64898/2026.09.08.26362398

**Authors:** Asma M. Aman, Eslam Zaher, Santiago Diaz-Torres, Sjoerd J. Driessen, Victor A. de Vries, Frank CT. van der Heide, Antonia Kolovos, Joshua M. Schmidt, Henry N. Marshall, Lania Saleh, Alicia Schulze, Gabriëlla A. Blokland, Carroll A. Webers, Carla J. van der Kallen, Anke Wesselius, Ilja Arts, Freekje van Asten, Mathias Gorski, Martina E. Zimmermann, Klaus J. Stark, Iris M. Heid, Terri L. Young, Louis R. Pasquale, Ayellet V. Segrè, Janey L. Wiggs, Anthony P. Khawaja, Donald J. Zack, Raymond C. Wong, Alex W. Hewitt, Alexander K. Schuster, Tos T. Berendschot, Alberta A. Thiadens, Karin A. van Garderen, Caroline C.W. Klaver, Pirro G. Hysi, Christopher J. Hammond, Caroline Brandl, Jamie E. Craig, Wishal D. Ramdas, Ya Xing Wang, Jost B. Jonas, Fred Roosta, Michael L. Hunter, Stuart MacGregor, Samantha S. Lee, David A. Mackey, Maciej Trzaskowski, Puya Gharahkhani

## Abstract

The peripapillary retinal nerve fibre layer (pRNFL) thickness and Bruch’s membrane opening-minimum rim width (BMO-MRW) are three-dimensional retinal biomarkers for glaucoma. We aimed to demonstrate that AI-derived thickness from two-dimensional fundus images can act as proxies, enabling the discovery of novel neurodegenerative loci. AI-derived pRNFL and BMO-MRW strongly correlated with OCT-derived thickness (r: 0.69 and 0.79, respectively). After validation, these phenotypes were predicted in two cohorts lacking disc-centred OCT: the UK Biobank and the Canadian Longitudinal Study on Aging. The predicted phenotypes showed strong genetic correlations with directly measured phenotypes from a previous study for both phenotypes (0.70 and 0.96, respectively). This data increased statistical power, identifying 29 loci for pRNFL thickness and 122 loci for BMO-MRW, including 14 loci that were independent of VCDR. We observed shared and sector-specific thickness loci overlapping glaucoma loci and revealed loci independent of known risk factors. Together, these results emphasise that multidimensional inferences can be drawn from 2D imaging, enabling downstream genetic analyses.

## Background

Primary open-angle glaucoma (POAG) is a leading cause of irreversible blindness worldwide, characterised by progressive retinal ganglion cell loss and optic nerve damage^1,2^. This damage can be quantified by structural parameters derived from optical coherence tomography (OCT) scans: peripapillary retinal nerve fibre layer (pRNFL) thickness and Bruch’s membrane opening-minimum rim width (BMO-MRW)^3^.

Current POAG management focuses on controlling intraocular pressure (IOP); however, a substantial proportion of patients continue to experience glaucoma progression despite well-controlled IOP. POAG can also occur within the normal range of IOP (< 21 mmHg) in a proportion of individuals, indicating the presence of IOP-independent disease mechanisms. All loci identified by glaucoma genome-wide association studies (GWAS) to date are associated with either IOP or vertical cup-to-disc ratio (VCDR)^4,5^, where VCDR is a quantitative measure of optic nerve integrity which is often used as a proxy of the neurodegenerative component in glaucoma. Expanding the known genetic architecture of the optic nerve is likely feasible by analysing BMO-MRW, which provides anatomically accurate and precise assessment of neuroretinal rim tissue and optic neuropathy^6^, and ganglion cell-related layer estimates in the papillary region, which are highly relevant to glaucoma.

GWAS is used to identify genetic variants associated with complex traits, often requiring large sample sizes to identify associated variants with small effect sizes. Given that it is challenging to obtain sufficient statistical power to test the association between loci and the trait, AI has been increasingly used to predict clinical phenotypes from medical imaging and other clinical data to improve phenotyping in large datasets. For example, a previous study predicted VCDR from fundus images, increasing power for genetic discovery compared to manually labelled values^4^. Several large-scale biobanks lack optic disc-centred OCT scans needed to measure pRNFL thickness and BMO-MRW, but they do contain thousands of 2D retinal fundus images. As such, this presents an opportunity to leverage AI models to infer 3D OCT-derived values from 2D fundus images and further leverage this approach for downstream genetic discovery.

GWAS meta-analyses have been conducted previously on pRNFL thickness and BMO-MRW, but their small sample sizes had limited potential for genetic discovery^7^. By using AI-derived pRNFL thickness and BMO-MRW, we expanded the sample size to conduct the largest GWAS meta-analyses for both phenotypes to date. Here, we show that AI-derived pRNFL thickness and BMO-MRW from 2D fundus images can act as proxies for the 3D measures in two large biobanks (UK Biobank and Canadian Longitudinal Study of Aging, CLSA), enabling the discovery of novel risk loci to study the associated underlying biology of glaucoma (**Figure 1**).

**Figure 1.**
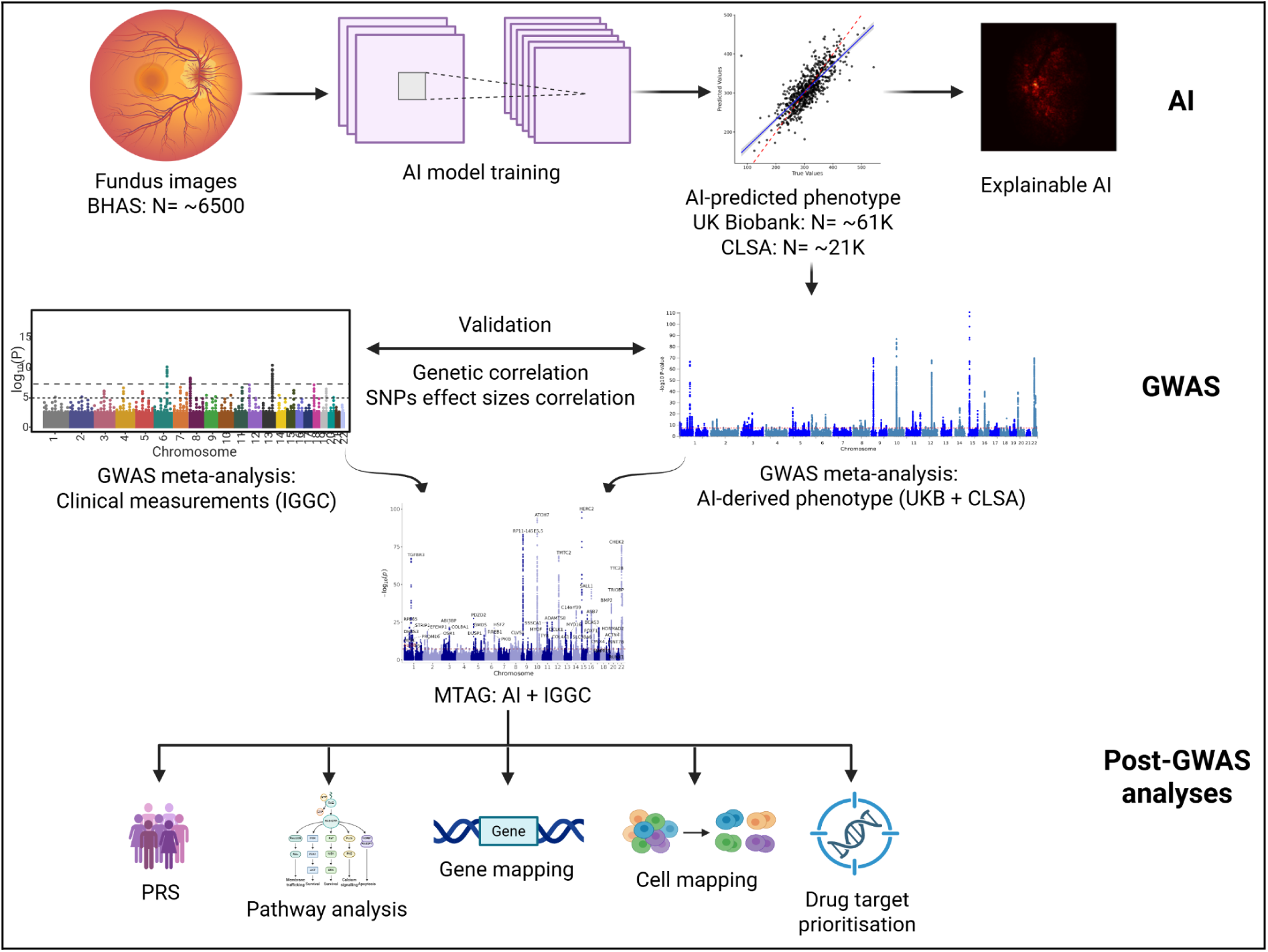
Overview of the study design and analysis pipeline. The figure summarises the three main stages of this study. The same pipeline was conducted for each phenotype included in this work. (1) AI framework: convolutional neural network-based modelling, phenotype generation in the UK Biobank and Canadian Longitudinal Study of Aging (CLSA) cohorts, and explainable AI. (2) GWAS analyses: genome-wide association analyses for the AI-derived phenotype, validation using directly measured thickness GWAS results previously reported from the International Glaucoma Genetics Consortium (IGGC), and integration of AI-derived and IGGC GWAS using MTAG analysis. (3) Post-GWAS analyses: gene mapping, pathway enrichment analysis, spatial cell-type mapping, drug target prioritisation, and polygenic risk scores (PRS) construction. This figure was created in BioRender. Diaz, S. (2026) https://BioRender.com/fabxn0s.

## Methods

### Phenotypes

Definitions of global and sectoral pRNFL thickness and BMO-MRW have been described previously^7^. In brief, pRNFL contains the axons of the retinal ganglion cells and the thickness represents the area surrounding the optic nerve head, while BMO-MRW is the minimum distance between the Bruch’s membrane opening and the inner limiting membrane, quantifying the thickness of the neuroretinal rim. The optic nerve head is typically divided into six sectors (i.e., temporal, superotemporal, inferotemporal, nasal, superonasal, and inferonasal) by OCT scans to facilitate analysis of both phenotypes. Global values for both thickness measurements refer to the weighted average of their respective sectors.

## Datasets

### UK Biobank

This is a large population-based cohort comprising approximately half a million participants aged between 40 and 69 recruited from the United Kingdom between 2006 and 2010^8^. The study includes extensive genetic and health-related data. Among the full cohort, approximately 68,000 participants underwent fundus photography and OCT imaging at baseline, and ∼19,000 participants were imaged during the first repeat assessment visit, with around 2000 participants overlapping between both visits. Fundus photographs and OCT images were acquired using the Topcon 3D OCT-1000 Mark II system; this provided retinal thickness centred around the macular region but does not allow estimation of thicknesses near the optic nerve head. The UK Biobank study received ethical approval from the North West Multi-centre Research Ethics Committee, and all participants provided written informed consent.

### The Canadian Longitudinal Study on Aging (CLSA)

CLSA is a large-scale longitudinal cohort that enrolled individuals aged between 45 and 85 and collected extensive genetic and phenotypic data at baseline^9,10^. The study contains approximately 57,377 fundus photographs acquired using Topcon (TRC-NW8) non-mydriatic retinal camera at baseline. The images, covering both left and right eyes, were obtained from around 30,000 participants. Notably, OCT data were not collected. Ethics approval was obtained from the relevant institutional review boards of all participating institutions.

### International Glaucoma Genetics Consortium (IGGC)

The consortium includes multiple cohorts from various countries, focusing on glaucoma and its endophenotypes. GWAS meta-analyses conducted by IGGC identified 9 loci associated with global pRNFL thickness using data from 25,942 participants. Additionally, 9 loci associated with global BMO-MRW were identified in analyses of 12,080 participants. The analyses further revealed 28 loci for sectoral pRNFL thickness and 19 loci for sectoral BMO-MRW. All participants included in these analyses were of European ancestry. Detailed descriptions of GWAS design, quality control, and meta-analysis procedures are provided in the original IGGC paper^7^.

### The Busselton Healthy Ageing Study (BHAS)

The BHAS is a community-based cohort study that is part of the IGGC, where we used imaging data to train AI models to predict retinal thicknesses. The study was conducted in the south-west of Western Australia and recruited 5,029 adults born between 1946 and 1964. All participants underwent comprehensive eye examinations, including tonometry and fundoscopy. Among these participants, 3,574 participants had pRNFL thickness and/or BMO-MRW measurements obtained using Spectralis OCT scan with Eye Explorer software v5.7.5.0 (Heidelberg Engineering, Heidelberg, Germany). pRNFL thickness was measured using a 3.45 mm diameter peripapillary circular scan manually centred on the optic nerve head (12°, 768 A-scans, 100 automated real-time [ART] averages), while BMO-MRW was quantified using 24 radial scans spaced 7.5° apart. At baseline, 2.1% (75/3563) of participants who had pRNFL thickness and/or BMO-MRW self-reported a diagnosis of glaucoma. The study received ethics approval from the Human Research Ethics Committee in the University of Western Australia (Number RA/4/1/2203)^11,12^.

### The Beijing Eye Study (BES)

BES is a population-based cohort study conducted in northern China^13–15^. The baseline examination was performed in 2001, with follow-up examinations in 2006 and 2011. In 2011, a total of 3,468 participants aged 50 years or older underwent comprehensive ocular examinations, including OCT imaging (Spectralis®, wavelength: 870 nm; Heidelberg Engineering, Heidelberg, Germany). pRNFL thickness was measured using a circular B-scan centred on the optic disc (3.5 mm diameter), consisting of 100 single A-scans. Genetic data were collected from 1,646 participants. The study protocol was approved by the Ethics Committee of Beijing Tongren Hospital, and all participants provided written informed consent.

### Artificial Intelligence approaches Image quality assessment model

Training AI models requires good-quality images to extract the important features and reduce noise. Given the large number of fundus images in the datasets, we developed a model to assess the quality of images instead of assessing them manually. We used ResNet-18, a convolutional neural network model, which was pre-trained on ImageNet (i.e., a dataset containing around 15 million general images). The model was fine-tuned using 1,298 fundus images (649 bad, 649 good) obtained from the UK Biobank. The image quality labels were derived from previous clinician grading^16^ and divided into 80% training and 20% validation. Afterwards, the model was tested using 198 images (100 good and 98 bad) from an independent dataset (1000-image dataset^17^), where the image quality labels were assessed manually based on general criteria including adequate brightness and contrast, absence of blur, and clear optic disc features. We reported performance metrics including accuracy, precision, and recall.

### Developing thickness prediction model

We utilised the EfficientNet_V2_L model pre-trained on ImageNet attached to a Bayesian classifier. The models were fine-tuned using around 6,500 high-quality fundus images, including both eyes, which were selected using the image quality assessment model, as well as global and sectoral pRNFL thickness and BMO-MRW labels obtained from BHAS. Sectoral thickness measurements are more susceptible to extreme values than global measurements, which represent average thickness values across sectors. Accordingly, outliers were identified and removed from sectoral data using the interquantile range (IQR) method (Q1 - 4 × IQR, Q3 + 4 × IQR). The images were divided into three sets: 72% for the training set, 18% for the validation set, and 10% for the test set. During training, images were resized to 256×256 pixels and augmented using random rotation (5 degrees), horizontal flipping (P = 0.5), random brightness adjustment (P = 0.3), and Gaussian blurring (P = 0.1), followed by tensor conversion and normalisation. Validation images were only resized and normalised without augmentation.

The training was initiated by unfreezing the classifier layer, while keeping the hidden layers frozen. Once the classifier was trained, the hidden layers were unfrozen to allow for fine-tuning. The model was trained using a mean square error loss function, with an additional constraint that penalised predictions outside a realistic thickness range defined as six standard deviations from the mean. To mitigate overfitting, we applied the early stopping method which terminates the training once the validation loss shows no improvement over a specified number of epochs. We predicted pRNFL thickness and BMO-MRW in the test set and reported Pearson’s correlation coefficient (r), root mean square error (RMSE), mean absolute error (MAE), and coefficients of determination R^2^. PyTorch (v2.2.1) was used to develop the model.

### Model explainability

We performed model explainability analyses for our thickness models trained on the 2D fundus images. In this setting, the goal was to assess whether our models made predictions based on valid plausible features of the input that concentrate on anatomically relevant regions for pRNFL thickness and BMO-MRW rather than spurious image cues. We employed Manifold Integrated Gradients (MIG)^18^ to generate saliency maps for each fundus image. These maps assign an importance score to each pixel, indicating how strongly it contributes to the model’s predicted thickness value. Visualising these scores as a heatmap provides an interpretable summary of the image evidence the network uses, allowing us to verify that predictions are driven by anatomically relevant structures.

MIG is a perceptually robust variant of Integrated Gradients^19^, which uses a generative vision model to capture a latent representation of the data manifold and generates explanations that are faithful to both the model and data used for training. The resulting saliency maps are more robust, stable, and perceptually smoother.

### Genome-wide association studies

#### Discovery

The trained model was used to predict global and sectoral pRNFL thickness and BMO-MRW measurements from fundus images in the UK Biobank and CLSA. We then conducted GWAS using the predicted values for each phenotype from the UK Biobank and CLSA using Regenie^20^ (v2.2.4). GWASs were adjusted for age, sex, and 10 principal components. To account for eye size, we included refractive error as a covariate in the UK Biobank as a proxy for axial length. In addition, we adjusted CLSA GWAS summary statistics for refractive error using refractive error GWAS meta-analysis of European ancestry with 542,934 participants using conditional analysis through the mtCOJO method^21,22^ (v1.91.7 beta1), as refractive error and axial length data were not available in CLSA. We then meta-analysed the results from the UK Biobank (N = 61,258 for global pRNFL thickness, global BMO-MRW, and sectoral BMO-MRW; N = 61,255 for sectoral pRNFL) and CLSA (N = 21,888 and N = 21,887 for the corresponding measures, respectively).

### Replication

The IGGC GWAS meta-analysis, which is based on actual OCT-derived measurements, was used as a replication dataset, and the correlation of the independent genome-wide significant SNPs’ effect sizes was measured. In addition, we estimated the genetic correlation using the bivariate LD score regression (LDSC) method^23,24^ (v1.0.1) between AI-based and IGGC GWAS meta-analyses.

### AI and IGGC MTAG

We combined the UK Biobank and CLSA GWAS meta-analysis with the IGGC GWAS meta-analysis to improve the statistical power. To mitigate heterogeneity, we integrated the results using multi-trait analysis of GWAS (MTAG) (v1.0.8)^25^. We calculated SNP heritability using the univariate LDSC method. Independent genome-wide significant SNPs were identified using LD clumping in the FUMA platform^26^, with a genome-wide significance threshold of p1 = 5 × 10^-8^, an LD clumping threshold of r^2^ = 0.2, and a clumping window of 1 Mb. The most significant SNP within a locus was designated as the lead SNP. To account for multiple testing across sectors, we further corrected the significance threshold of sectoral pRNFL and BMO-MRW by dividing it by the number of sectors (P = 5 × 10^-8^ / 12). Global measures were not included in this correction because they represent composite measures derived from the sectoral phenotypes. Using the mtCOJO method, we conditioned the summary statistics of global BMO-MRW on VCDR to find VCDR-independent loci.

### Post-GWAS analyses

#### Gene-based association analysis

We employed the mBAT-combo^27,28^ method (v1.94.1) to find gene-trait associations for global pRNFL thickness and BMO-MRW. This method is particularly robust when multiple SNPs within a gene have masking effects. The gene-level significance threshold was determined by accounting for all evaluated genes and traits using the Bonferroni method: P < 1.3×10^-6^ (0.05 / (18,992 genes × 2 traits tested)).

### Pathway enrichment analysis

Pathway analysis was performed to identify biological pathways enriched for gene-level associations for global pRNFL thickness and BMO-MRW. MAGMA^29^ gene set analysis was employed to conduct this analysis on the FUMA platform^26^. Predefined gene sets were obtained from the Molecular Signatures Database (MsigDB v2023.1Hs). Pathway significance was determined using Bonferroni correction across all evaluated pathways and traits; P < 1.5×10^-6^ (0.05/ (17,023 gene sets × 2 traits tested)).

### Glaucoma loci independent from IOP and VCDR

We performed two MTAG analyses of POAG, one incorporating global pRNFL thickness and the other incorporating global BMO-MRW to boost the power of our GWAS to identify glaucoma loci which are driven by pRNFL thickness or BMO-MRW. Summary statistics for POAG were obtained from a European POAG GWAS using the following datasets: UK Biobank, CLSA, IGGC, and Mass General Brigham Biobank (N_Cases_ = 29,241; N_Controls_ = 350,181). This POAG GWAS identified 86 loci. Any locus that was genome-wide significant in the MTAG analysis and did not reach nominal significance (P ≥ 0.05) for either IOP or VCDR was considered independent of these phenotypes. Loci with suggestive association (P < 0.005) with one or both phenotypes were considered potentially independent. IOP summary statistics were obtained from a GWAS meta-analysis of 133,492 participants^30^, while VCDR summary statistics were derived from a GWAS meta-analysis of 111,724 individuals, in which VCDR was estimated from AI-based labelling of fundus images and adjusted for vertical disc diameter^4^.

### Summary-based-data Mendelian randomisation (SMR)

To identify potential causal genes underlying global pRNFL thickness and BMO-MRW, we applied the SMR^31^ method (v1.3.1), integrating GWAS summary statistics with quantitative trait loci (xQTL). The analyses were implemented in the SMR portal (https://yanglab.westlake.edu.cn/smr-portal/)^32^. We leveraged blood- and brain-derived multi-omics data, including expression QTL (eQTL) data from the eQTLGen consortium (N = 31,684; N_genes_ = 11,106) and BrainMeta (N = 2,865; N_genes_ = 10,520), splicing QTL (sQTL) from GTEx v8 (N = 755; N_genes_ = 1,918) and BrainMeta (N = 2,865; N_genes_ = 9,465), methylation QTL (mQTL) from McRae (N = 1,980; N_genes_ = 13,142) and BrainMeta (N = 1,160; N_genes_ = 14,588), as well as blood-derived proteome QTL (pQTL) obtained from the FENLAND study (N = 10,708; N_genes_ = 1,540), INTERVAL study (N = 3,301; N_genes_ = 614), and SCALLOP study (N = 30,931; N_genes_ = 71).

A heterogeneity in dependent instruments (HEIDI) test^31^ was performed to assess potential pleiotropy, using a significance threshold of P_HEIDI_ > 0.05. Genes were considered significant if the SMR P-value passed multiple testing correction using the Bonferroni method (P < 0.05 / (number of genes in xQTL × 2)) and showed no evidence of heterogeneity in the HEIDI test.

### Gene prioritisation

We prioritised genes for drug-gene interaction evaluation by selecting genes that were significant in at least one tissue and supported by at least three lines of genetic evidence, including mBAT-combo, SMR analyses incorporating eQTL, sQTL, pQTL, and/or mQTL. In addition, we utilised the drug-gene interaction database (https://dgidb.org/) to identify drug-gene interactions and potential repurposing drug candidates for neuroprotection.

### Spatial cell-type mapping

We performed spatial mapping of cells for global pRNFL thickness and BMO-MRW using the gsMap (v1.71.2) (genetically informed spatial mapping of cells for complex traits) method^33^. GsMap integrates GWAS summary statistics with spatial transcriptomics data to identify cell populations enriched for genetic risk of complex traits. In this method, genetic specificity scores (GSS) are measured, which quantifies how specifically a gene is expressed in a spot (i.e., the spatial unit) relative to other spots. These scores are then used to annotate SNPs and assess trait enrichment using stratified LDSC.

GWAS summary statistics for global pRNFL thickness and BMO-MRW were used. Spatial transcriptomics data was obtained from a publicly available dataset of human retina samples obtained from healthy adult donors^34^. Quality control (QC) and preprocessing were performed using Scanpy and pandas packages in Python 3.11. Raw count matrices were integrated with spot-level metadata indexed by spatial barcodes. Barcode concordance and per-column missingness were assessed as QC metrics, and a unified annotation field (obs["annotation"]), derived from labels2)) was created with missing labels assigned as "Unknown". No additional gene or unique molecular identifier (UMI) level filtering was applied.

After QC, the number of spots per donor was as follows: Donor2 (N = 669), Donor3 (N = 869), Donor4 (N = 1,417), and Donor4_s2 (N = 2,181). Among these, Donor2 and Donor4_s2 contained the largest number of spots annotated in the optic nerve region, which is the most relevant anatomical region for pRNFL thickness and BMO-MRW. Therefore, these two donors were selected for gsMap to ensure sufficient representation of the optic nerve. The final datasets comprised of 669 spots for Donor 2 (Choroid/Sclera: 274; Optic nerve: 160; Retina: 59; Unknown: 176), and 2,181 spots for Donor4_s2 (Choroid/Sclera: 1,334; Optic nerve: 522; Retina: 215; Unknown: 110).

### Polygenic risk scores

Polygenic risk scores (PRS) were constructed for global pRNFL thickness using GWAS summary statistics and the Bayesian regression framework implemented in SBayesRC^35^ (v0.2.5). SBayesRC integrates LD information with functional annotations to estimate posterior SNP effect sizes. The LD reference panel was obtained from European participants from the UK Biobank, which was provided by the developers. The resulting posterior effect sizes were used as weights to compute PRS in BES (N = 1,046) using PLINK (v2). PRS was standardised before downstream analysis. The association between PRS and pRNFL thickness was assessed using a linear regression model adjusted for covariates such as age, sex, axial length, genotyping batch, and the first 5 principal components. Predictive performance was evaluated by comparing models with and without PRS and calculating incremental R^2^.

## Results

### AI

#### Images quality assessment

The ResNet-18 image quality assessment model achieved an accuracy of 0.91, with a precision of 0.93 and a recall of 0.88 in the internal validation set. On the external test set (1000-image dataset), the model achieved an accuracy of 0.96, with a precision of 0.96 and a recall of 0.97. Subsequently, the model was applied to large-scale datasets, where we obtained 131,791 high-quality fundus images for both eyes from the UK Biobank’s first and second instances, 47,764 from CLSA, and 9,766 from BHAS.

#### Thickness prediction

The global pRNFL thickness was normalised using a mean of 95.91 μm and a standard deviation (SD) of 11.85. The mean of predicted pRNFL thickness in the test set was 95.44 μm, and the SD was 7.65. In the test set, AI-predicted pRNFL thickness showed strong correlation with clinically measured values (r = 0.69; **Supplementary Figure 1-a**). The model demonstrated moderate predictive performance with R^2^ = 0.48, with MAE = 6.23 μm and RMSE = 8.80 μm. For global BMO-MRW, the measurements were standardised using a mean of 309.91 μm and an SD of 58.46. The mean of predicted BMO-MRW was 310.96 μm, and the SD was 49.61. Predicted and clinically measured BMO-MRW showed strong correlation (r = 0.79; **Supplementary Figure 1-b**). The model explained 63% of the variance (R^2^ = 0.63), with an MAE of 25.36 μm and an RMSE of 34.42 μm.

The prediction performance was comparable across pRNFL sectors, with the nasal and temporal regions having the lowest explained variance (R^2^ = 0.43) and inferonasal having the highest R^2^ of 0.52 (**Supplementary Table 1**). For sectoral BMO-MRW, the range was between 0.5 and 0.57 for temporal and superonasal, respectively (**Supplementary Table 2**).

### Model explainability

#### Global Models

To examine the spatial features driving each model’s predictions, saliency maps were generated using MIG for randomly selected fundus images for the models trained to estimate global pRNFL thickness and BMO-MRW (**Figure 2**). Each set of input images is paired with its corresponding saliency maps, indicating the regions that most strongly influenced the predicted thickness. Despite the absence of any spatial supervision or segmentation guidance during training, both models exhibited consistent and anatomically coherent attention patterns that aligned closely with the structural basis of their respective OCT-derived targets.

**Figure 2.**
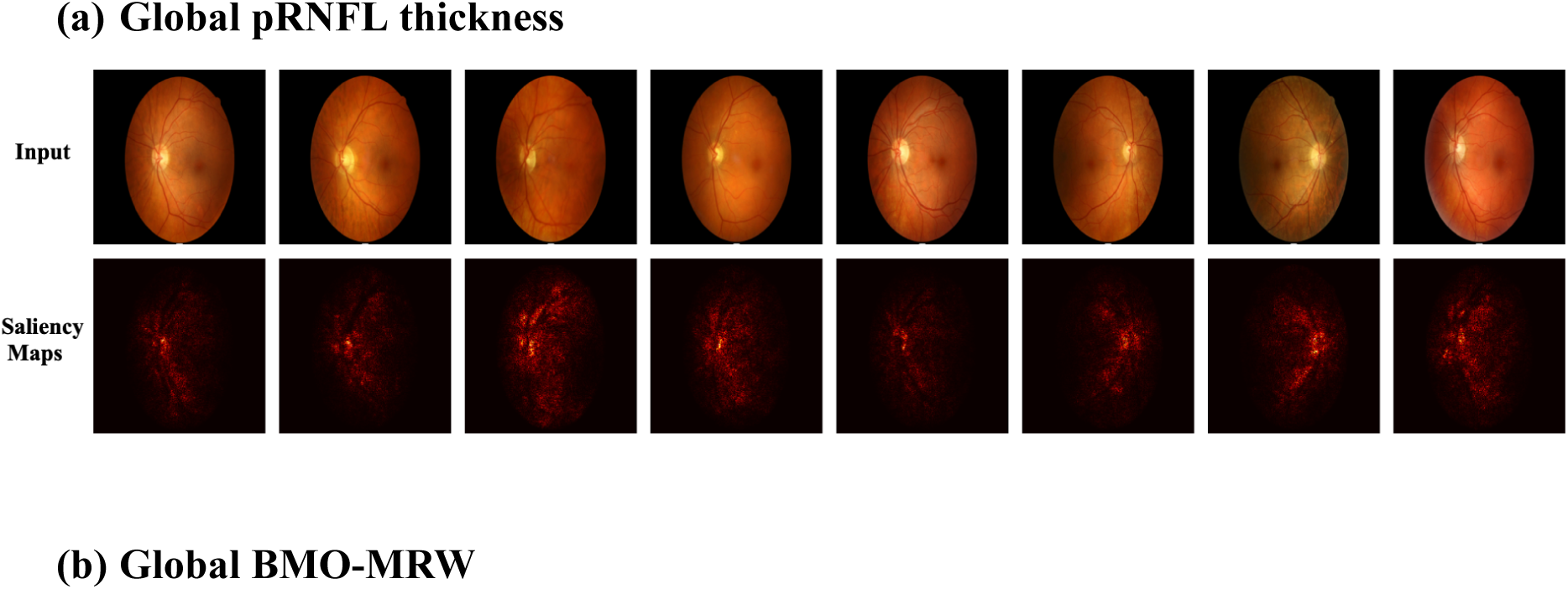
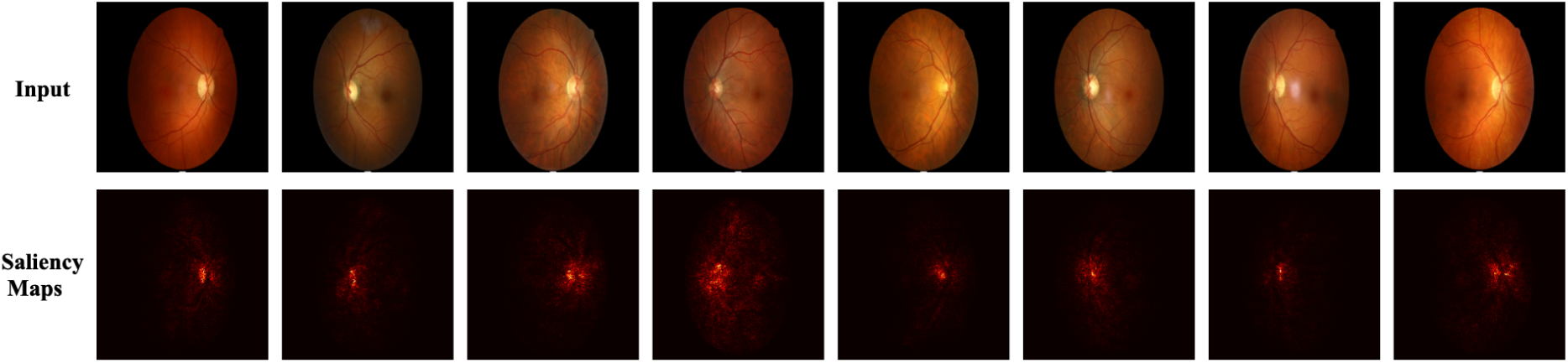
Saliency-based model explainability for global thickness prediction. (a) Global pRNFL thickness and (b) Global BMO–MRW models. For each panel, the first row shows randomly sampled input fundus images, and the second row shows their corresponding saliency maps indicating the image regions most influential for the predicted thickness. Consistent with the distinct anatomical domains of the two thicknesses, pRNFL saliency is predominantly distributed in the peripapillary region surrounding the optic disc, whereas BMO–MRW saliency is more tightly localised to the optic nerve head and neuroretinal rim.

In the pRNFL model (**Figure 2-a**), the saliency responses were broadly distributed around the optic disc, forming a smooth peripapillary ring that corresponds to the anatomical course of the retinal nerve fibre layer. The model appeared to rely on features within this circumferential region, reflecting the spatial domain over which RNFL thickness is conventionally defined. In contrast, the BMO–MRW model (**Figure 2-b**) showed a markedly more confined activation pattern, with attributions concentrated within the optic nerve head and peaking along the neuroretinal rim at the disc margin.

The distinct localisation of attention in these two models underscores their alignment with complementary structural correlates of the optic nerve head. The pRNFL network emphasises the peripapillary retinal tissue surrounding the disc, whereas the BMO–MRW network attends to the inner rim architecture itself. Together, these results indicate that the models have learned spatially specific, anatomically meaningful representations consistent with the geometry of their respective reference measurements, supporting the interpretability and biological plausibility of their predictions.

### Sectoral Models

**Supplementary Figure 2** extended the saliency analysis to the sectoral models and showed a consistent spatial shift in attributions as the target region moved around the optic disc. For the sectoral pRNFL models (**Supplementary Figure 2-a**), saliency remained predominantly peripapillary, with the temporal model emphasising regions lateral to the disc, the superotemporal model concentrating in the upper-temporal quadrant, and the inferotemporal model concentrating in the lower-temporal quadrant; similarly, nasal-sector saliency appeared on the nasal side of the disc, while the superonasal and inferonasal models showed more spatially concentrated emphasis in the upper-nasal and lower-nasal peripapillary regions, respectively. For the sectoral BMO-MRW models (**Supplementary Figure 2-b**), attributions were more disc-centred across sectors, with sectoral variation expressed primarily as shifts in rim-localised emphasis in the corresponding quadrants rather than extension into the surrounding peripapillary retina. Notably, the nasal pRNFL sector exhibited comparatively more diffuse and fragmented saliency relative to the other sectoral models, suggesting less localisation for that target in the fundus images.

### GWAS

#### Discovery and replication

The LDSC analysis showed that the AI-derived global pRNFL thickness GWAS meta-analysis exhibited modest SNP-based heritability (h^2^_SNP_ = 0.13, SE = 0.01). Mild genomic inflation was observed (λ_GC_ = 1.17), while the LDSC intercept was 1.01 (SE = 0.01), suggesting that the inflation was largely attributable to polygenicity rather than confounding. The AI-derived global BMO-MRW meta-analysis showed modest SNP-based heritability (h^2^_SNP_ = 0.27, SE = 0.02), with mild genomic inflation (λ_GC_ = 1.27) and an LDSC intercept of 1.04 (SE = 0.01). Q-Q plots for global and sectoral phenotypes are shown in **Supplementary Figure 3,** together with the corresponding LDSC output for each sector. Bivariate LDSC analysis of the AI and OCT-derived GWAS meta-analyses revealed strong and statistically significant genetic correlation for global pRNFL thickness (rg = 0.70, SE = 0.07, P = 2 × 10^-25^) and global BMO-MRW (rg = 0.96, SE = 0.06, P = 5 × 10^-68^). Genetic correlation estimates for sector-specific pRNFL varied across sectors, with the strongest correlation observed for the inferonasal region (rg = 0.86, SE = 0.07, P = 1.52 × 10^-37^) and the weakest for the nasal region (rg = 0.37, SE = 0.09, P = 2.24 × 10^-5^), as shown in **Supplementary Table 3**. This is consistent with the qualitative explainability patterns in **Supplementary Figure 2**, where the nasal pRNFL sector exhibited more fragmented saliency, in line with its weaker genetic correlation estimate compared with other regions. In contrast, genetic correlations were constantly very strong across BMO-MRW sectors in the range between 0.87 and 0.99 (**Supplementary Table 4**).

The AI-derived GWAS meta-analysis identified 22 genome-wide significant loci for global pRNFL thickness (**Supplementary Table 5**) and 97 loci for global BMO-MRW (**Supplementary Table 6**). Additionally, the total number of independent loci across pRNFL sectors was 61 after correcting for the number of sectors (P < 4.2 × 10^-9^) (**Supplementary Table 7**), of which inferonasal and superonasal analyses identified the highest number of loci with 28 and 26, respectively. Also, the total number of independent loci across BMO-MRW sectors was 101 (P < 4.2 × 10^-9^) (**Supplementary Table 8**), of which superonasal and inferonasal analyses also revealed the highest number of loci across BMO-MRW sectors with 74 and 71 loci, respectively. Manhattan plots for global phenotypes are presented in **Figure 3**, while sectoral plots are shown in **Supplementary Figure 4**.

**Figure 3.**
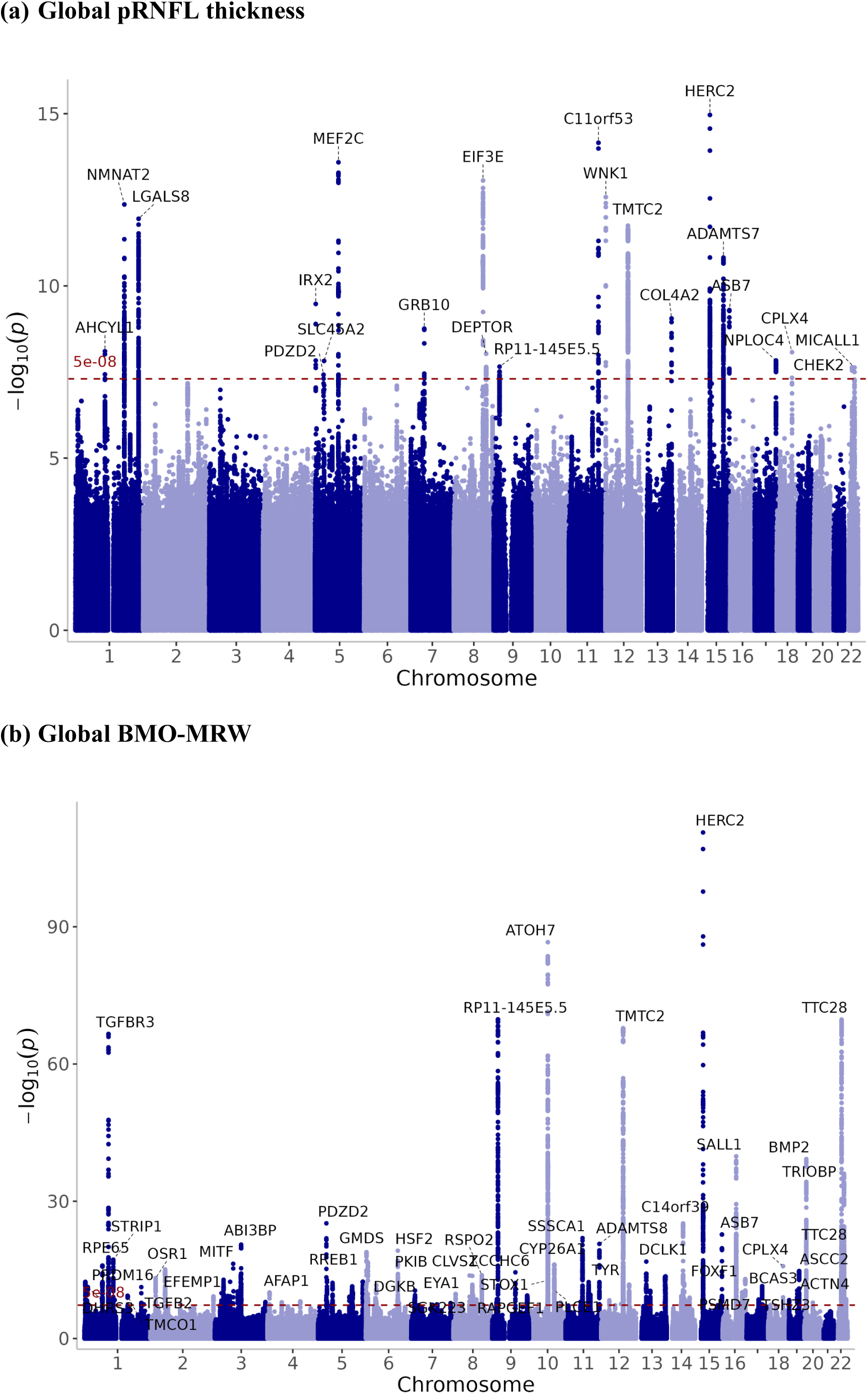
Manhattan plots for AI-derived GWAS meta-analysis. (a) Global pRNFL thickness and (b) Global BMO-MRW. The −log10(P-value) of SNP associations is plotted against genomic position across chromosomes. The dashed red horizontal line represents the genome-wide significance threshold (P = 5 × 10^-8^). Each dot represents one SNP. Genome-wide significant loci were defined using LD clumping (r^2^ < 0.2 within 1 Mb).

The top genome-wide significant SNPs’ effect size correlation was 0.62 (P = 0.005) and 0.82 (P = 2.2 × 10^-24^) for global pRNFL thickness and global BMO-MRW, respectively. The scatter plots are shown in **Figure 4**. For sectoral pRNFL, the strongest correlation was for inferonasal with an estimate of 0.86 (P = 3.1 × 10^-11^), while the lowest was for the superotemporal sector with a value of 0.51 (P = 0.042) **(Supplementary Figure 5-a)**. On the other hand, the correlations across BMO-MRW sectors were all strong **(Supplementary Figure 5-b)**.

**Figure 4.**
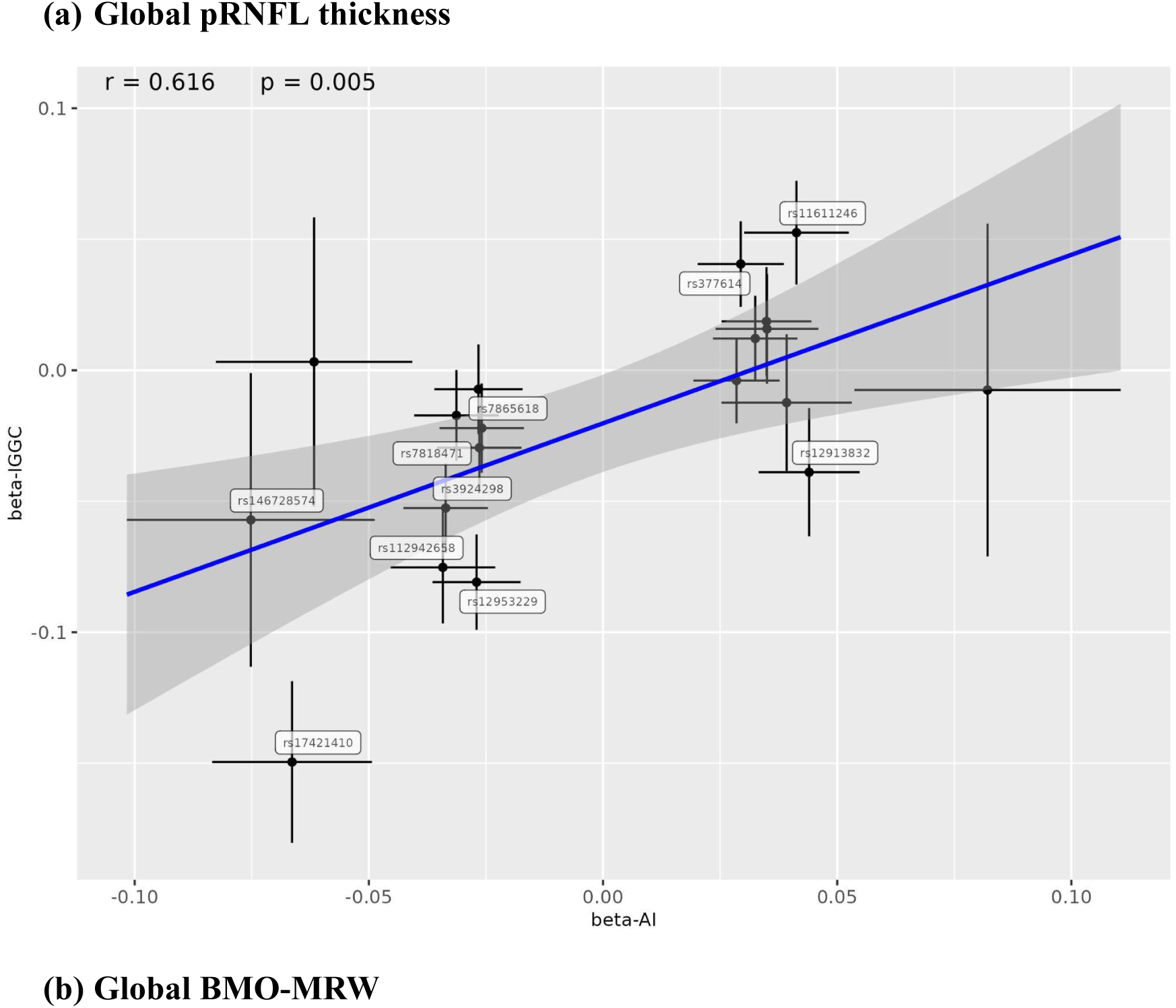
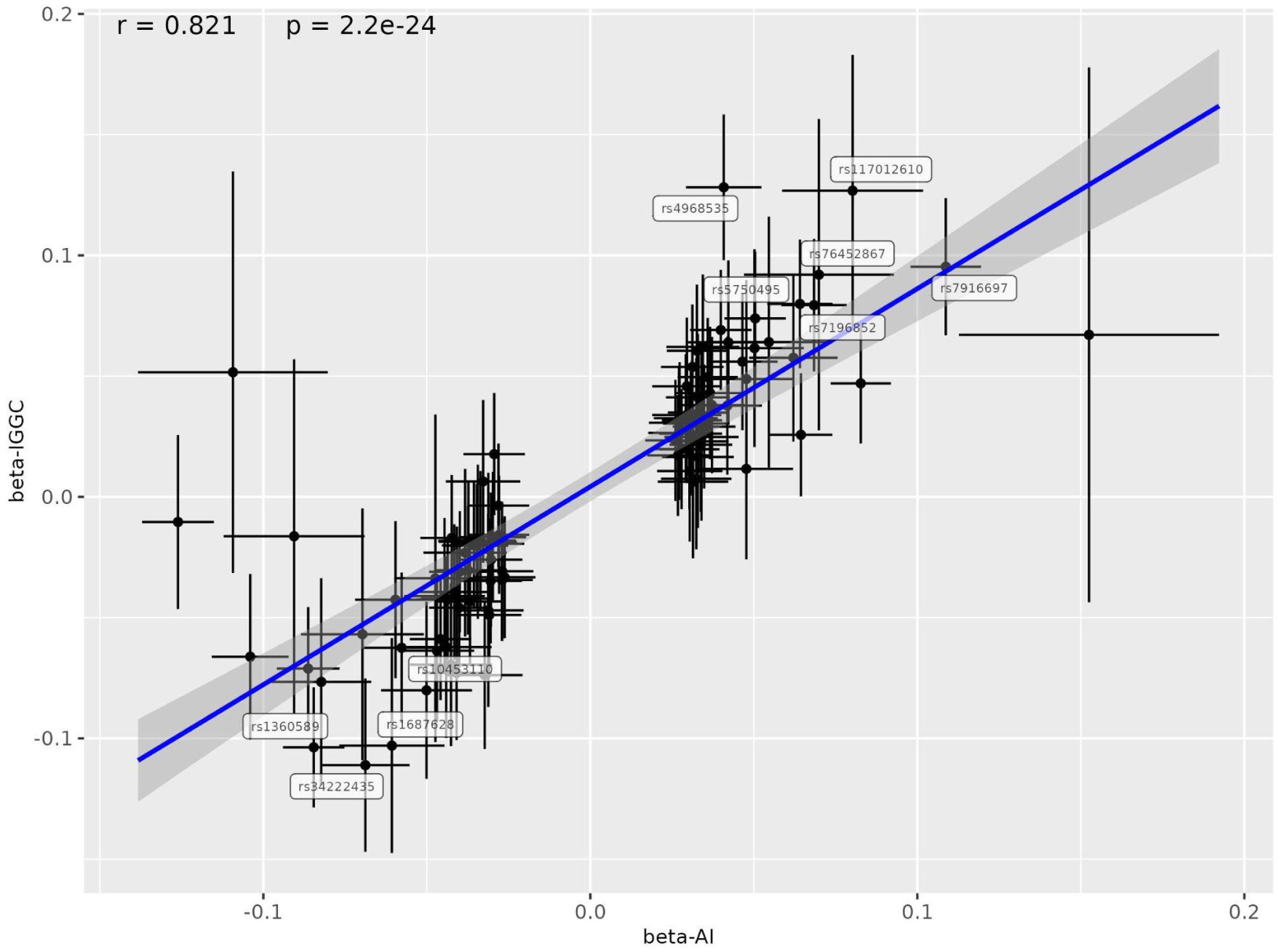
Concordance of SNP effect sizes between AI-derived and directly measured thickness GWAS. (a) Global pRNFL thickness and (b) Global BMO-MRW. Scatter plots compare effect size estimates (β) for lead SNPs at genome-wide significant loci (P < 5 × 10^-8^) identified in the AI-derived GWAS (x-axis) and in GWAS of directly measured thickness in IGGC cohorts (y-axis). Horizontal and vertical error bars around each lead SNP represent 95% confidence intervals for effect size estimates in the AI-derived and IGGC GWAS, respectively. The blue line indicates the linear regression fit. Each point represents one lead SNP. Pearson’s correlation coefficient (r) and P values are shown.

### AI and IGGC GWAS MTAG

#### Global pRNFL thickness and BMO-MRW

After combining AI and IGGC results using the MTAG method, 29 significant loci were identified for pRNFL thickness (**Figure 5-a; Supplementary Table 9**). Among them, there were 9 known glaucoma loci such as rs17421410 (near *TMEM161B*), rs10811644 (near *CDKN2B*), and rs12913832 (near *HERC2*).

**Figure 5.**
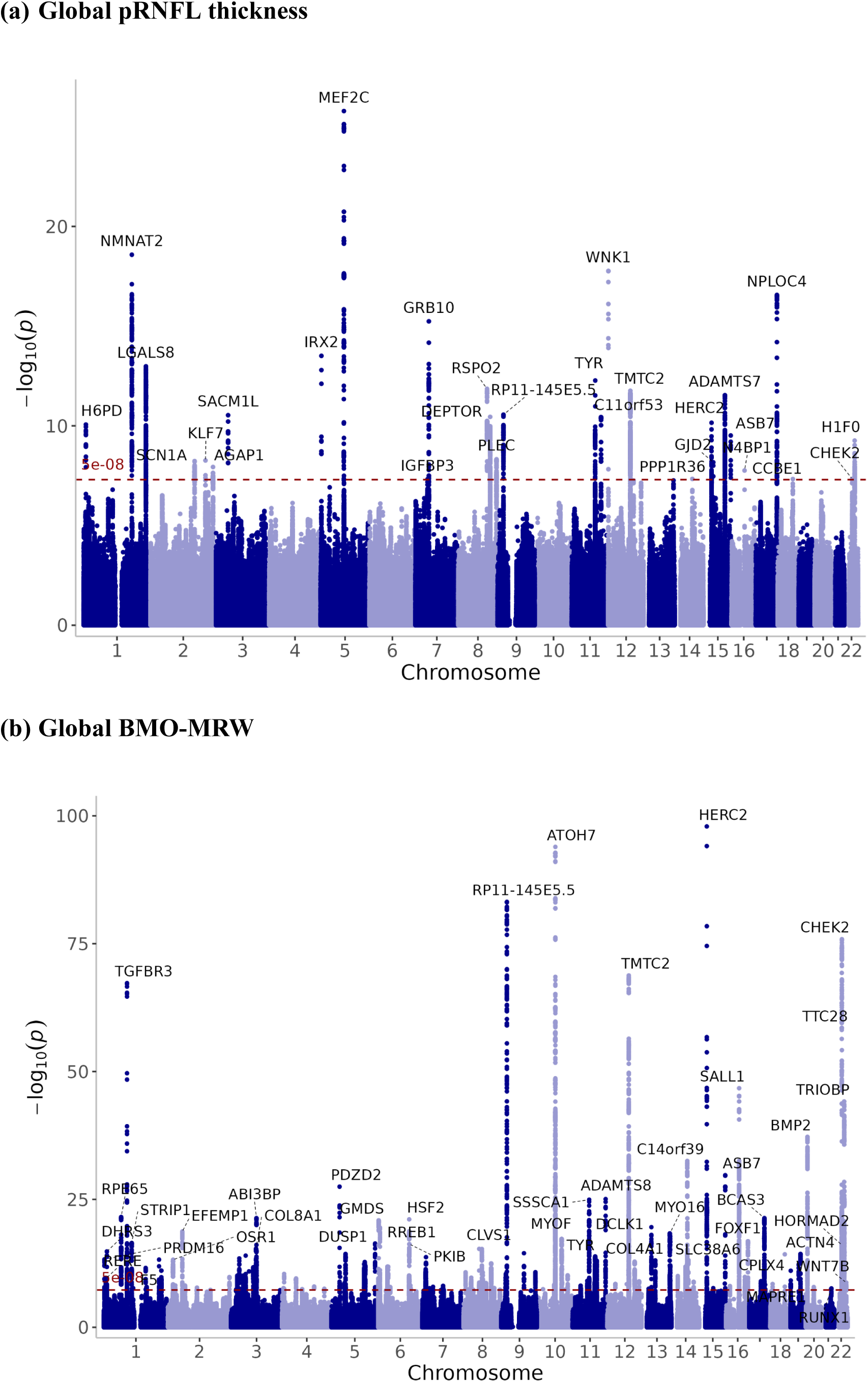
Manhattan plots for AI-derived and directly measured thickness MTAG analysis. (a) Global pRNFL thickness and (b) Global BMO-MRW. The −log10(P-value) of SNP associations is plotted against genomic position across chromosomes. The dashed red horizontal line represents the genome-wide significance threshold (P = 5 × 10^-8^). Each dot represents one SNP. Genome-wide significant loci were defined using LD clumping (r^2^ < 0.2 within 1Mb).

For BMO-MRW AI and IGGC MTAG results, we found 122 significant loci, of which 14 loci remained significant after adjusting for VCDR using the mtCOJO method, demonstrating VCDR-independent effects (**Figure 5-b; Supplementary Table 10**). In total, there were 64 glaucoma loci associated with global BMO-MRW, such as rs1192419 (near *HFM1*), rs1126809 (near *TYR*), rs12913832 (near *HERC2*), and rs5756813 (near *H1-0*).

### Sectoral pRNFL and BMO-MRW

Sectoral pRNFL analyses identified a total of 79 loci (P < 4.2 × 10^-9^), of which 13 loci were associated previously with glaucoma (**Supplementary Table 11**). We observed both broadly shared association patterns and regionally enriched effects across sectors at glaucoma-associated loci. For example, rs17421627 (near *TMEM161B*) was identified as a shared locus across sectors; however, effect sizes were at least two-fold larger in nasal regions compared to temporal regions. Also, the locus near *SIX6* demonstrated associations across all sectors but with opposite effect size directions between temporal and nasal regions (P = 2.43 × 10^-40^ for effect size difference between temporal and nasal; P = 1.26 × 10^-50^ between inferior regions; and P = 2.43 × 10^-18^ between superior regions). Superonasal and inferonasal exhibited the largest effect sizes across several glaucoma loci, including rs5008216 (near *FMNL2*) and rs10120688 (near *CDKN2B*). The variant rs6117293 (near *BMP2*) exhibited its strongest association in the inferonasal region (β = 0.028).

On the other side, sectoral BMO-MRW analyses identified in total 112 loci, of which 69 loci were associated previously with glaucoma (**Supplementary Table 12**). In general, loci showed mainly similar effect sizes across sectors, with a few exceptions such as rs12913832 (near *HERC2*), where temporal, inferotemporal, and inferonasal exhibited substantially larger effect sizes (β = −0.12, −0.14, and −0.19, respectively).

### Post-GWAS analyses

#### Gene-based association analysis

The mBAT-combo analysis identified 21 independent loci encompassing 62 genes significantly associated with pRNFL thickness after Bonferroni correction (P < 1.3 × 10^-6^) (**Supplementary Table 13**). Three loci were novel compared to the pRNFL thickness per-SNP GWAS. *TMEM161B*, *CDKN2B* and *TRIOBP*, known glaucoma genes, were associated with pRNFL thickness. Similarly, the analysis revealed 375 significant genes (P < 1.3×10^-6^) within 82 independent loci in association with BMO-MRW, of which 8 loci were additional over the GWAS loci (**Supplementary Table 14**). Several known glaucoma genes were identified, such as *CDKN2B*, *SIX6*, and *TMCO1*.

### Pathway-based enrichment analysis

MAGMA gene-set enrichment analysis highlighted REACTOME_MELANIN_BIOSYNTHESIS and GOBP_HINDLIMB_MORPHOGENESISv pathways as significantly associated with pRNFL thickness (P_Bonferroni_ < 0.05; **Supplementary Table 15**). The results suggest roles for pigmentation and developmental processes in pRNFL thickness. Furthermore, MAGMA revealed 25 pathways enriched for BMO-MRW (P_Bonferroni_ < 0.05; **Supplementary Table 16**). These pathways spanned diverse biological processes, including pigmentation, morphogenesis, and developmental processes, among others, indicating broad biological involvement in BMO-MRW.

### Glaucoma loci independent of IOP and VCDR

The European MTAG analysis of POAG and global pRNFL thickness revealed 94 loci associated with POAG (**Supplementary Table 17**). Among these, rs58265464 (near *HMGXB3*)^36,37^ was independent from IOP and VCDR (both P > 0.05) and associated with primary angle-closure glaucoma, while rs4390362 (near *B3GAT1*)^5^, and rs2836855 (near *GET1*)^38^ were potentially independent (P_IOP and/ or P_VCDR > 0.005).

The MTAG analysis with global BMO-MRW identified 125 loci associated with POAG (**Supplementary Table 18**). Among these, the SNP rs58265464 (near *HMGXB3*)^36,37^ was also independent from IOP and VCDR. Five loci were potentially independent including rs2443722 (near *VGLL4*), rs3909355 (near ENSG00000229855)^5^, rs12421242 (near *YAP1*)^5^, rs4842316 (near *PAWR*)^37^, and rs12886636 (near *TTLL5*)^5^. Notably, these independent loci were revealed in larger cross-ancestry POAG GWAS, suggesting that pRNFL thickness and BMO-MRW can help identify additional mechanisms for glaucoma, as most of the currently identified loci are IOP and/or VCDR loci.

### SMR and genes prioritisation

SMR analyses followed by HEIDI tests for global pRNFL thickness identified 7 unique potential causal genes with supporting evidence from eQTL, 3 genes from sQTL, 10 from mQTL, and none survived the significance criteria for pQTL. For global BMO-MRW, the analyses revealed 32 unique potential causal genes with supporting evidence from eQTL, 13 from sQTL, 3 from pQTL, and 56 from mQTL. The complete list of genes identified by SMR xQTL analyses for global pRNFL thickness and BMO-MRW can be found in **Supplementary Tables 19-20**, respectively.

Three genes were prioritised as potential drug target candidates for global pRNFL thickness by three lines of evidence, including mBAT-combo and SMR analysis with supporting evidence from multiple QTLs. The prioritised genes were *LGALS8*, *PLEC*, and *TRIOBP*. On the other hand, 6 genes were prioritised for global BMO-MRW by at least three pieces of evidence, including *CTSW*, *FLNB*, *HAUS4*, *LTBP3*, *NDUFAF3*, and *P4HA2*. **Table 1** lists all prioritised genes for global pRNFL thickness and BMO-MRW. **Supplementary Tables 21-22** show genes prioritised by two lines of genetic evidence. Drug-gene interaction screening using the Drug-Gene Interaction platform identified metformin hydrochloride, which is a widely used drug for type 2 diabetes, as an interactor of *NDUFAF3*.

**Table 1.** List of prioritised genes supported by three lines of genetic evidence. (a) Global pRNFL thickness and (b) Global BMO-MRW. Gene prioritisation was performed by selecting those that were significant in at least one tissue and three lines of genetic evidence, including mBAT-combo, SMR analyses incorporating either eQTL, sQTL, pQTL, and/or mQTL.

**(a) Global pRNFL thickness**
| Gene | CHR | Evidence types | Function |
| --- | --- | --- | --- |
| *LGALS8* | 1 | mBAT-combo, mQTL-Blood, mQTL-Brain, sQTL-Brain | Encodes galectin-8 which regulates cell adhesion, immune responses and inflammatory processes<sup>39,40</sup>. |
| *PLEC* | 8 | mBAT-combo, eQTL-Blood, eQTL-Brain, mQTL-Blood | Encodes plectin protein that links intermediate filaments to microtubules and other cytoskeletal components, helping maintain cytoskeleton structure and cellular integrity<sup>41</sup>. It was reported in association with Alzheimer disease<sup>42,43</sup>. |
| *TRIOBP* | 22 | mBAT-combo, eQTL-Brain, mQTL-Brain | Encodes multiple protein isoforms, which promote cytoskeletal organisation by stabilising and bundling actin filaments, contributing to cell structure and mechanical stability<sup>44,45</sup>. It was reported in association with POAG<sup>5</sup>, VCDR<sup>46</sup>, and IOP<sup>30</sup>. |

**(b) Global BMO-MRW**
| Gene | CHR | Evidence types | Function |
| --- | --- | --- | --- |
| *FLNB* | 3 | mBAT-combo, eQTL-Brain, sQTL-Brain | Encodes a protein that links actin filaments to membrane proteins, which enables signal transduction and regulates cell migration and adhesion<sup>47,48</sup>. It was reported in association with VCDR<sup>49</sup>, POAG<sup>5</sup>, and low-tension glaucoma<sup>38</sup>. |
| *NDUFAF3* | 3 | mBAT-combo, eQTL-Blood, mQTL-Brain | Encodes a mitochondrial complex I assembly factor, supporting oxidative phosphorylation and efficient energy production<sup>50</sup>. |
| *P4HA2* | 5 | mBAT-combo, mQTL-Blood, sQTL-Brain | Encodes a hydroxylase enzyme that is required for collagen biosynthesis and extracellular matrix remodelling<sup>51</sup>. It was reported for IOP<sup>52</sup>, VCDR<sup>49</sup>, and POAG<sup>37</sup>. |
| *CTSW* | 11 | mBAT-combo, eQTL-Blood, mQTL-Blood, mQTL-Brain | Encodes a cathepsin W, which is a protease enzyme that regulates aspects of T-cell functions<sup>53,54</sup>. |
| *LTBP3* | 11 | mBAT-combo, eQTL-Blood, mQTL-Blood | Regulates TGF-β signalling and secretion, contributing to extracellular matrix remodelling<sup>55</sup>. It was reported in association with POAG<sup>36</sup>, and VCDR<sup>49</sup>. |
| *HAUS4* | 14 | mBAT-combo, eQTL-Brain, mQTL-Blood, mQTL-Brain | Encodes a subunit of the augmin complex that contributes to mitotic spindle assembly and chromosome segregation<sup>56</sup>. It was reported for POAG<sup>37</sup> and retinal layer thickness<sup>57</sup>. |

### Spatial cell-type mapping

Using gsMap, global pRNFL thickness and BMO-MRW had significant enrichment in the optic nerve tissue. The top 100 genes contributing to the enrichment for global pRNFL thickness were predominantly ribosomal genes (*RPL* and *RPS* family members), as well as genes contributing to mitochondrial oxidative phosphorylation (e.g., *NDUFA4*), cytoskeletal organisation (e.g., *TUBA1B*), and immune-related processes (e.g., *HLA-B*) (**Supplementary Table 23**). This pattern suggests enrichment within metabolically active neuronal cells with high translational demand and energy production.

Among the top-ranked genes for BMO-MRW were established glaucoma-associated genes (e.g., *MYOC*, *CYP1B1*), extracellular matrix remodelling genes (e.g., *FBLN1*), regulators of transforming growth factor-β (TGF-β) (*LTBP* family members) (**Supplementary Table 24**). Immune-related genes within the HLA locus (e.g., *HLA-A*, *HLA-C*) were also observed among the top-ranked genes, together suggesting enrichment of tissue remodelling and immune-related pathways which are consistent with glaucoma pathophysiology.

### PRS

The standardised PRS result constructed from our global pRNFL thickness GWAS was significantly associated with global pRNFL thickness from the right eye (mean = 101.79 μm, SD = 12.07) in BES (N = 1,046; Females = 62.9%; Age: range = 50-93, mean = 63.5, SD = 9.3). In a linear regression model adjusted for age, sex, axial length, genotyping batch, and the first 5 principal components, each SD increase in PRS was associated with an increase in global pRNFL thickness (β = 1.80, SE = 0.36, P = 7.09 × 10^-7^). The PRS explained 2.1% of the phenotypic variance beyond covariates (incremental R^2^ = 0.021).

## Discussion

UK Biobank and CLSA are large datasets with over 525,000 participants together; however, both lack data on pRNFL thickness and BMO-MRW. In this study, we show that deep learning models can estimate 3D retinal thickness measurements from 2D fundus retinal images in these datasets, enabling large-scale phenotype generation for gene discovery while producing results consistent with those obtained from clinically estimated values. Using both phenotypes, we successfully identified neurodegenerative loci for glaucoma that are independent from known risk factors. These findings highlight the role of AI models in phenotyping and genetic discovery.

Our thickness prediction model with a Bayesian head achieved MAE comparable to or lower than previously reported MAE values, which range from 7.4 to 10.7 µm for pRNFL thickness prediction^58,59^ and 27.8 µm for BMO-MRW prediction^60^, demonstrating competitive prediction performance. Although the AI-derived values showed only moderate prediction accuracy with the clinical values, gene discovery using these estimated values was consistent with that obtained from clinically measured phenotypes with substantial overlap in the associated loci while enabling the identification of additional loci due to the larger sample size. In addition, the saliency maps demonstrated that the models relied on anatomically relevant regions to generate predictions, supporting the validity of those predicted estimates. Importantly, the primary aim for predicting the retinal thicknesses in this study was not to replace clinical measurements, but to produce proxy measurements that can increase the statistical power for gene discovery by enabling large-scale phenotype generation.

The majority of previously discovered POAG loci were associated with IOP and VCDR pathways^5^. Prior studies have suggested the presence of genetic mechanisms contributing to optic nerve vulnerability beyond IOP-mediated mechanisms^61^. By leveraging MTAG analysis of POAG with pRNFL thickness and BMO-MRW, we identified several POAG risk loci not associated with IOP and VCDR, suggesting potential mechanisms beyond these traditional risk factors. Given the substantially greater power of current POAG GWAS compared to the current GWAS of pRNFL thickness and BMO-MRW, the modest number of identified loci is expected. Future larger-scale pRNFL thickness and BMO-MRW GWASs may enable discovery of more glaucoma loci independent of IOP and VCDR and facilitate the identification of neuroprotective drug targets. Importantly, the number of loci identified through our MTAG analysis is not comparable to the 312 loci reported in the larger cross-ancestry multi-trait analysis of POAG^5^, as our analysis was restricted to European-only summary statistics and excluded the larger joint GWAS analysis incorporating IOP and VCDR.

The identification of both shared and sector-specific loci influencing pRNFL thickness relative to glaucoma risk highlights the complex genetic architecture of optic nerve vulnerability. Loci shared across sectors might reflect broader biological pathways related to eye development or generalised neurodegenerative processes. In contrast, sector-specific loci might indicate localised genetic effects leading to distinct patterns of pRNFL thinning in glaucomatous optic neuropathy observed in clinics^62–64^. Together, these findings suggest that genetic susceptibility of glaucoma partially involves region-specific mechanisms in the RNFL, highlighting the importance of considering genetics of pRNFL sectors in genetic analysis.

We prioritised three and six putative causal genes for global pRNFL thickness and BMO-MRW, respectively. Although several genes among them were previously associated with glaucoma, our multi-omics analysis provides strong evidence for their potential role as causal genes at these loci and linking retinal structural variation to glaucoma risk. Other genes that were not previously implicated in glaucoma may provide additional biological mechanisms for optic nerve vulnerability. The prioritised genes converge in biological pathways critical for glaucoma pathophysiology^65^, including immune modulation and neuroinflammation (*LGALS8*), cytoskeletal organisation (*PLEC*, *TRIOBP*), extracellular matrix remodelling (*FLNB*, *LTBP3*, *P4HA2*), cell cycle regulation and cellular maintenance (*HAUS4*), immune regulation (*CTSW*), and mitochondrial energy metabolism (*NDUFAF3*). Together, these findings support a role of optic nerve vulnerability mechanisms that extends beyond IOP-mediated pathways.

Metformin was revealed as a drug-gene interactor with *NDUFAF3*. As *NDUFAF3* is involved in mitochondrial complex I assembly, the observed interaction suggests that mitochondrial pathways may contribute to variation in pRNFL thickness, highlighting biological pathways that may influence retinal structural variation. The drug works by inhibiting the mitochondrial complex I, leading to activation of AMP-activated protein kinase (AMPK)^66^. Activation of AMPK may contribute to its protective effects against glaucoma, including reduction of IOP and anti-inflammatory actions that help protect retinal neurons from oxidative stress^67–71^. Metformin has been shown to protect retinal ganglion cells in a mouse model subjected to acute retinal ischemia resembling glaucomatous optic neuropathy^72^. Similarly, observational studies have reported an association between the use of metformin and lower risk of developing POAG among diabetic patients, suggesting a role in neuroprotection beyond glycemic control^73–75^. This is further supported by a recent Mendelian randomisation study that found potential protective effects of metformin on glaucoma risk^76^. These findings have led metformin to be evaluated as a potential therapeutic agent for POAG, as it is currently registered for a Phase II clinical trial (ClinicalTrials.gov ID: NCT05426044). This present study extends the work of predicting retinal thickness values from fundus images to evaluate the impact of the AI-derived values on gene discovery. Also, it represents the largest GWAS meta-analysis conducted to date for pRNFL thickness and BMO-MRW. However, several limitations should be noted. For instance, the analysis was restricted to participants of European descent because the AI models were trained on fundus images from European participants. As a result, the generalisability of the model to other ancestral groups remains uncertain. Future work should retrain and validate the model in diverse populations to ensure broader generalisability. In addition, the model was limited to older adults, reflecting the age distribution of the training data. Future model training should include participants from different age groups in order to predict their retinal thickness. Multi-omics data used for SMR analyses were obtained from brain tissue as a proxy for retinal tissue, as the latter has weak power to detect associations. Future studies leveraging larger retinal datasets will be important to capture retina-specific associations. Unlike pRNFL thickness, PRS analysis was not constructed for BMO-MRW, as data were not available in the target cohort.

In conclusion, we leveraged deep learning models to predict 3D retinal structural estimates from 2D fundus images, substantially increasing the sample size for genetic discovery. This approach revealed shared and sector-specific components of the genetic architecture underlying pRNFL thickness and BMO-MRW in relation to glaucoma risk and prioritised genes through multi-omics analysis. Our findings also identified additional genetic contributors to glaucoma risk related to retinal neurobiology that are independent of established risk factors. Together, these results advance the understanding of the genetic determinants of retinal structures and provide a framework for large-scale image-derived phenotyping and downstream genetic analyses.

## Contributors

A.M.A. participated in the study design, performed data analysis, and drafted the manuscript. E.Z. performed data analysis and drafted the manuscript. S.D.T. contributed to the data analysis and participated in the study design. P.G., M.T., and S.M. participated in the study design. P.G., M.T., D.A.M., and S.S.-Y.L. jointly supervised the research. S.S.-Y.L., S.J.D., V.A.d.V., F.C.v.d.H., A.K., J.M.S., H.N.M., L.S., A.S., G.A.B., C.A.W., C.J.v.d.K., A.W., I.A., F.v.A., M.G., M.E.Z., K.J.S., I.M.H, A.K.S., T.T.B., A.A.T., K.A.v.G., C.C.K., P.G.H., C.J.H., C.B., J.E.C., W.D.R., D.A.M, M.L.H., Y.W., J.B.J., D.J.Z., R.C.W., A.W.H. funding acquisition, data curation and/or providing summary statistics of individual GWAS. All authors revised and approved the final version of the manuscript.

## Acknowledgement

This research was made possible using the data and biospecimens collected by CLSA. This research has been conducted using the CLSA Baseline Comprehensive Dataset version 4.0 and baseline retinal images, under Application Number 190225. The CLSA is led by Drs. Parminder Raina, Christina Wolfson and Susan Kirkland. The authors gratefully acknowledge the time and commitment of the CLSA participants, without whom this research would not be possible. The opinions expressed in this manuscript are the author’s own and do not reflect the views of the Canadian Longitudinal Study on Aging. This study utilised data from the UK Biobank under application no. 25331. The authors would like to acknowledge the participants of the Busselton Health Study, the Raine Study, and their families for their ongoing participation. We thank the Raine Study team for their coordination and data collection, and we are grateful to the NHMRC and the Raine Medical Research Foundation for their long-term support over the past 30 years. We also acknowledge the ZIO Foundation (Vereniging Regionale HuisartsenZorg Heuvelland) for their contribution to The Maastricht Study. The researchers are indebted to all participants across the included cohorts for their willingness to take part in this research.

## Data availability

UK Biobank data are available through the UK Biobank Access Management System at (https://www.ukbiobank.ac.uk/). Data are available from the Canadian Longitudinal Study on Aging (www.clsa-elcv.ca) for researchers who meet the criteria for access to de-identified CLSA data. Data are available from The Maastricht, The AugUR, and ANZRAG Studies for any researcher who meets the criteria for access to confidential data; the corresponding author may be contacted to request data. Data described in the manuscript, code book, and analytic code will be made available upon request pending (e.g., application and approval, payment, other). TwinsUK welcomes data sharing with health researchers via a managed data access process which can be accessed at (https://twinsuk.ac.uk/researchers/access-data-and-samples/request-access/).

## Code availability

The analysis code used in this study is available upon request.

