## Supplementary Figures for "Genetic discovery using deep learning-derived optic nerve integrity phenotypes"

**Supplementary Figure 1. Scatter plot showing the relationship between true and predicted thickness values in the Busselton Healthy Aging Study.** (a) Global pRNFL thickness, (b) Global BMO-MRW, (c) Sectoral pRNFL thickness, and (d) Sectoral BMO-MRW. Each dot represents an individual sample. The solid blue line represents the linear regression fit with 95% confidence interval, while the dashed red line indicates the line of identity (perfect agreement). The Pearson's correlation coefficient (r) with the corresponding P value is displayed.

**(a) Global pRNFL thickness**

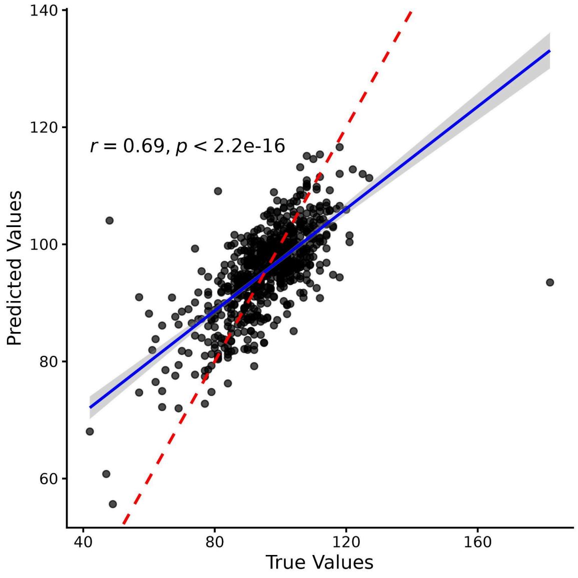


**(b) Global BMO-MRW**

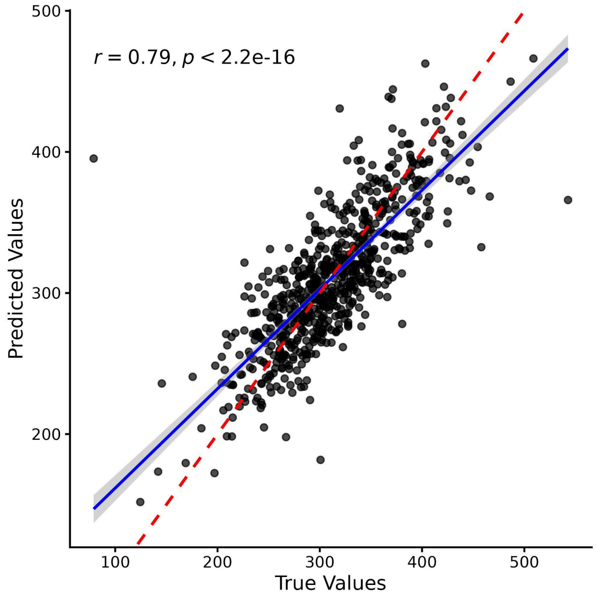

**(c) Sectoral pRNFL thickness**

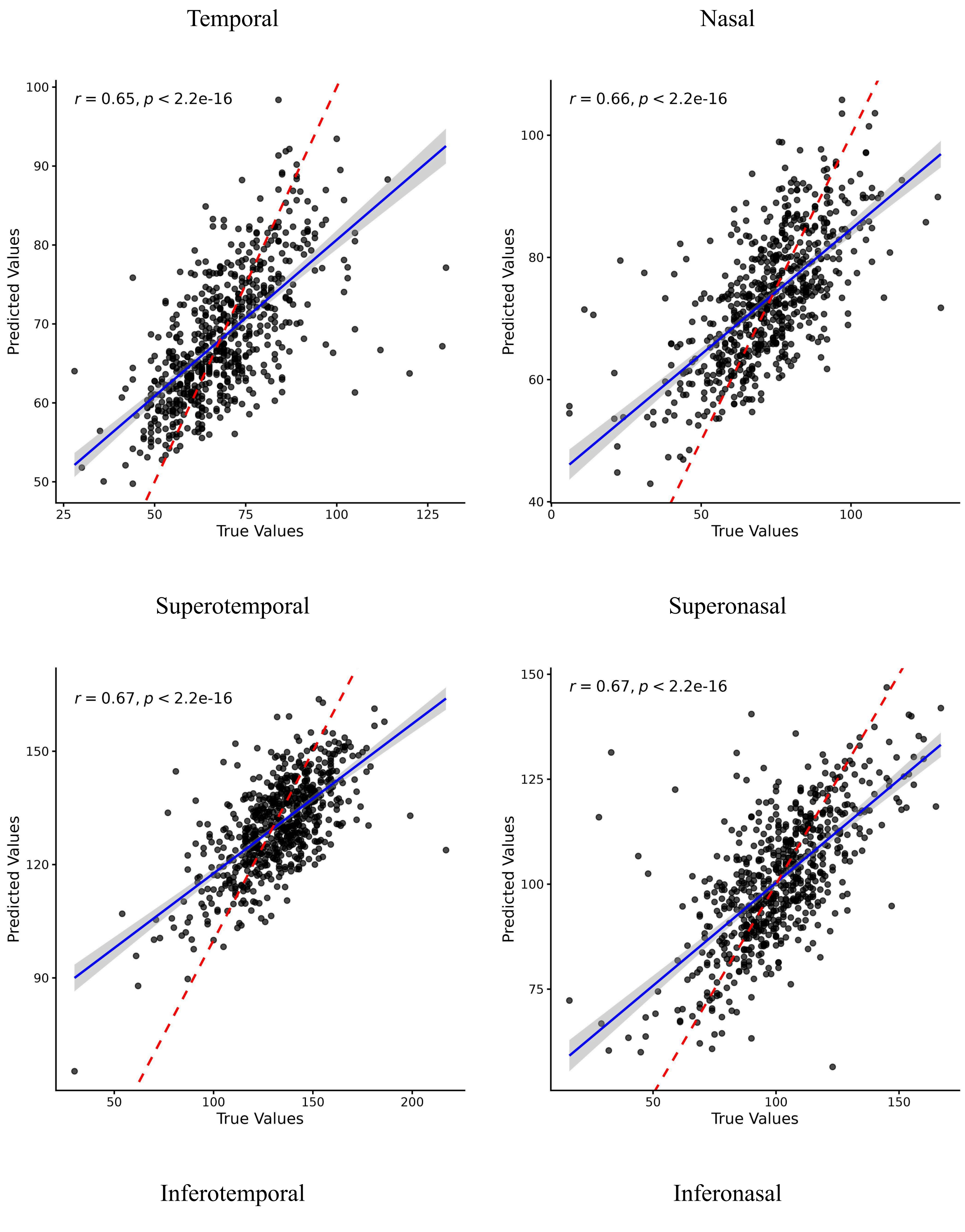

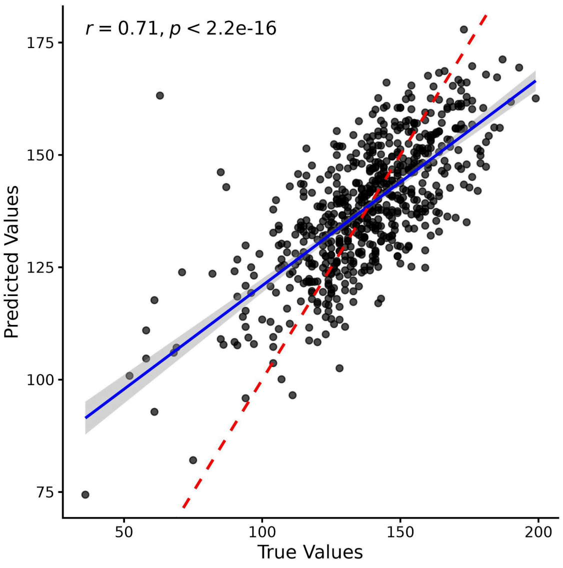


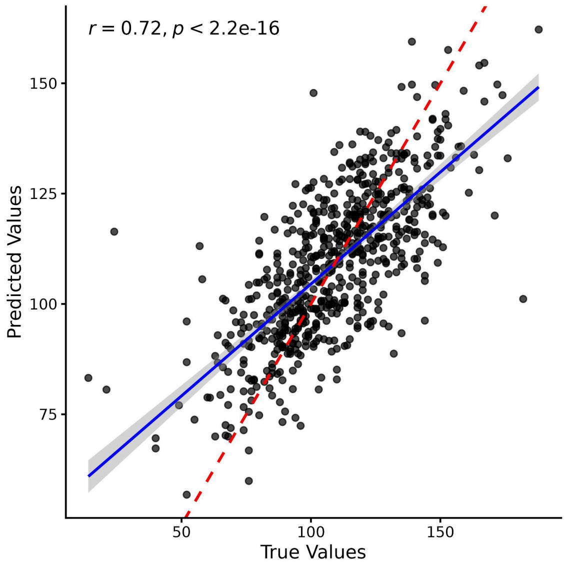


#### (d) Sectoral BMO-MRW

Temporal

Nasal

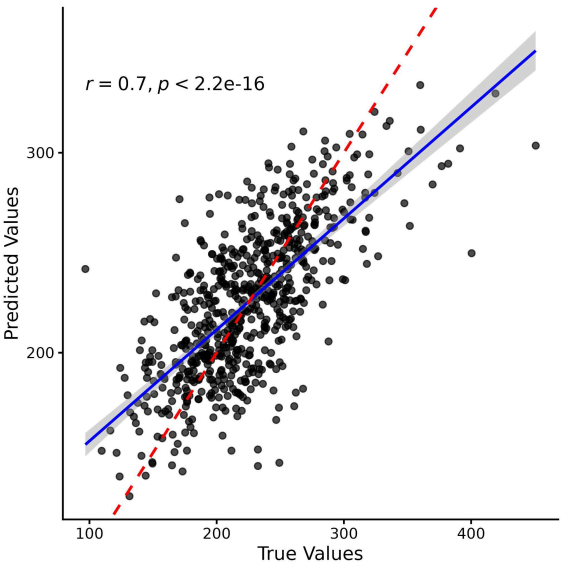


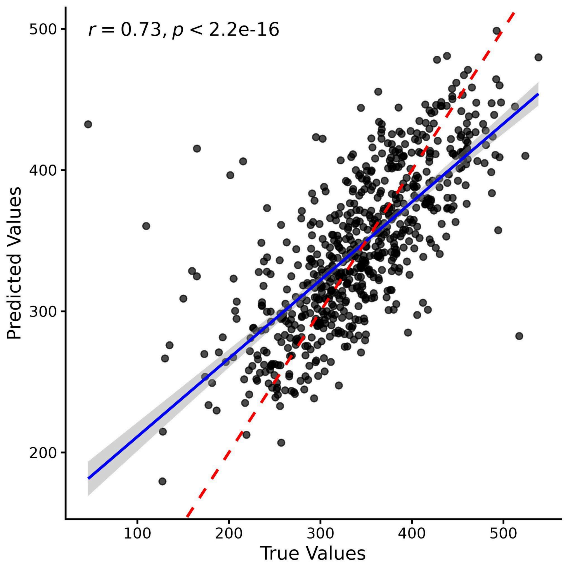


Superotemporal

Superonasal

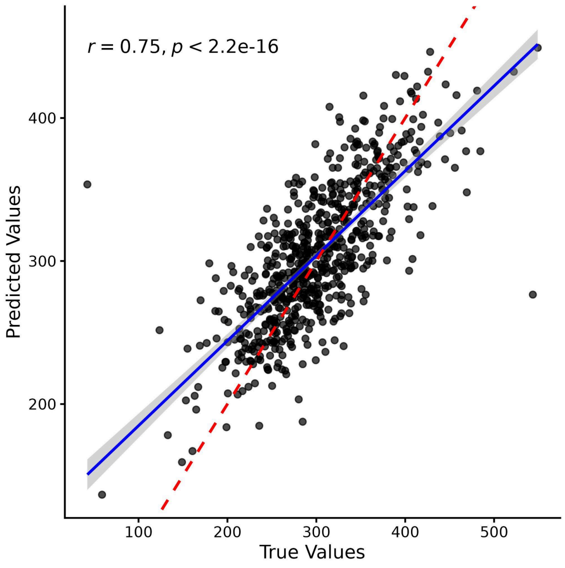


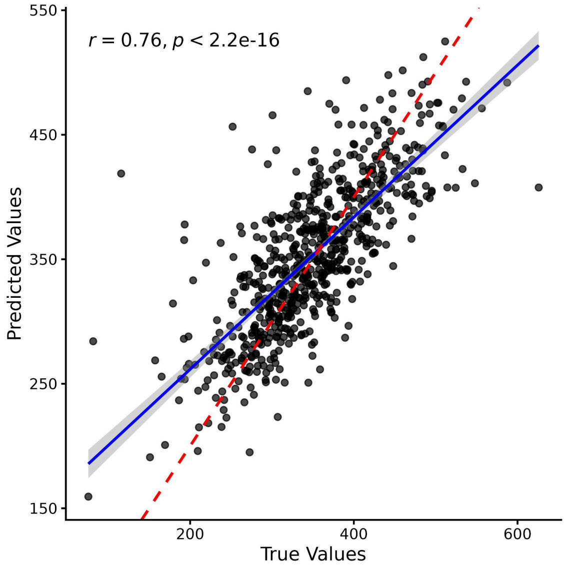


Inferotemporal

Inferonasal

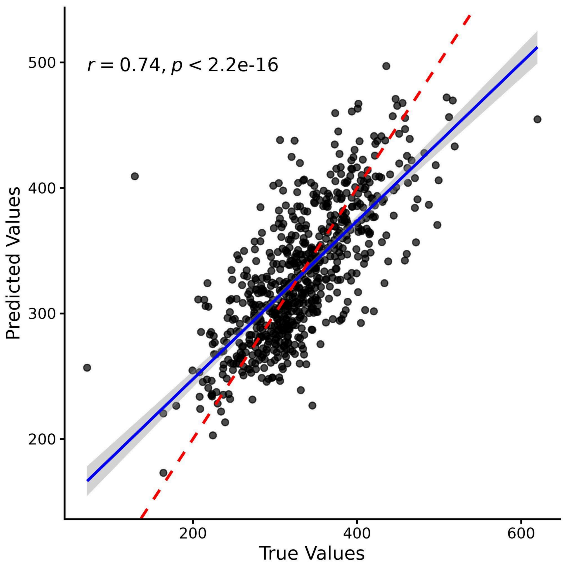


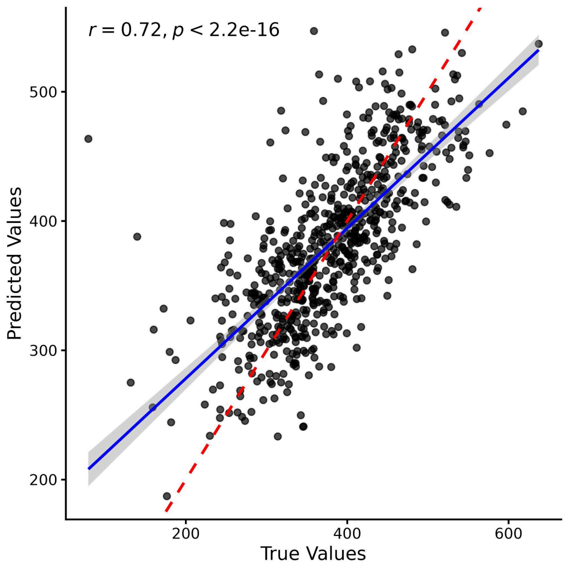

**Supplementary Figure 2. Saliency-based model explainability for sectoral retinal thickness prediction.** (a) Sectoral pRNFL thickness models and (b) sectoral BMO–MRW models, evaluated on the same set of input fundus images. For each panel, the first column shows the input fundus images and the subsequent columns show the corresponding saliency maps for the temporal, superotemporal, inferotemporal, nasal, superonasal, and inferonasal sectors. Across sectors, pRNFL saliency remains predominantly peripapillary and shifts around the optic disc in a sector-consistent manner, whereas BMO-MRW saliency is more disc-centred with sectoral variation expressed primarily as rim-localised emphasis.

**(a) Sectoral pRNFL thickness**

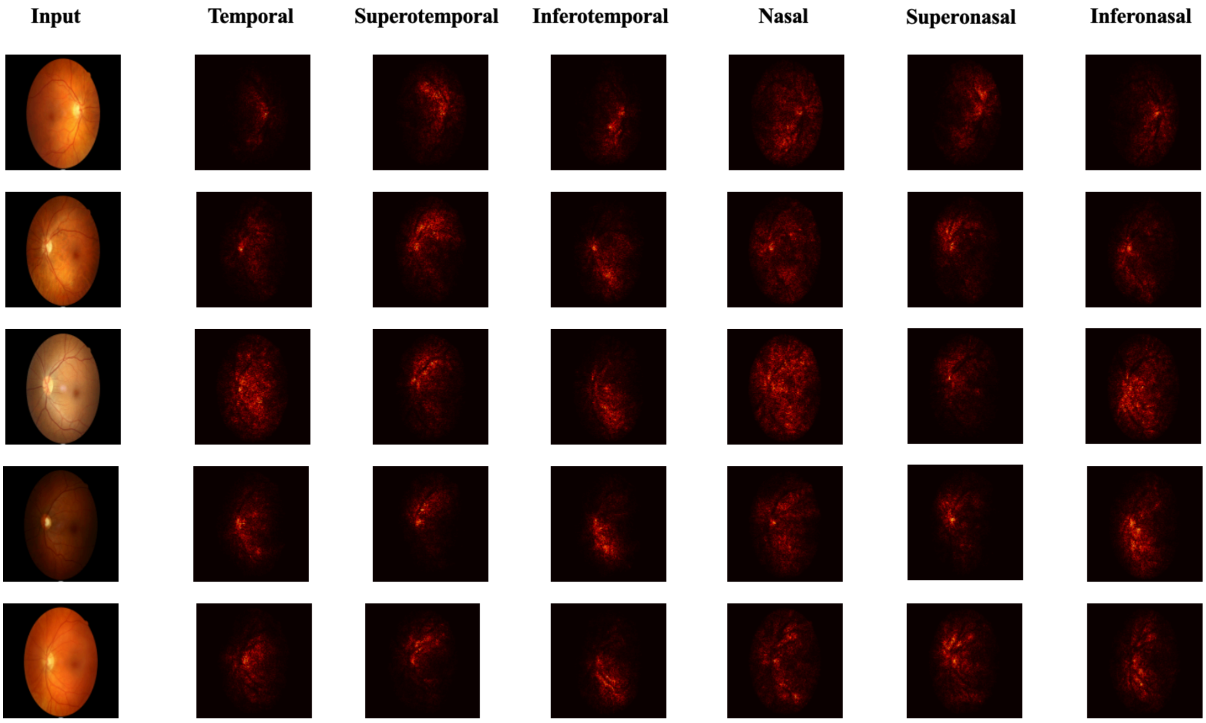


**(b) Sectoral BMO-MRW**

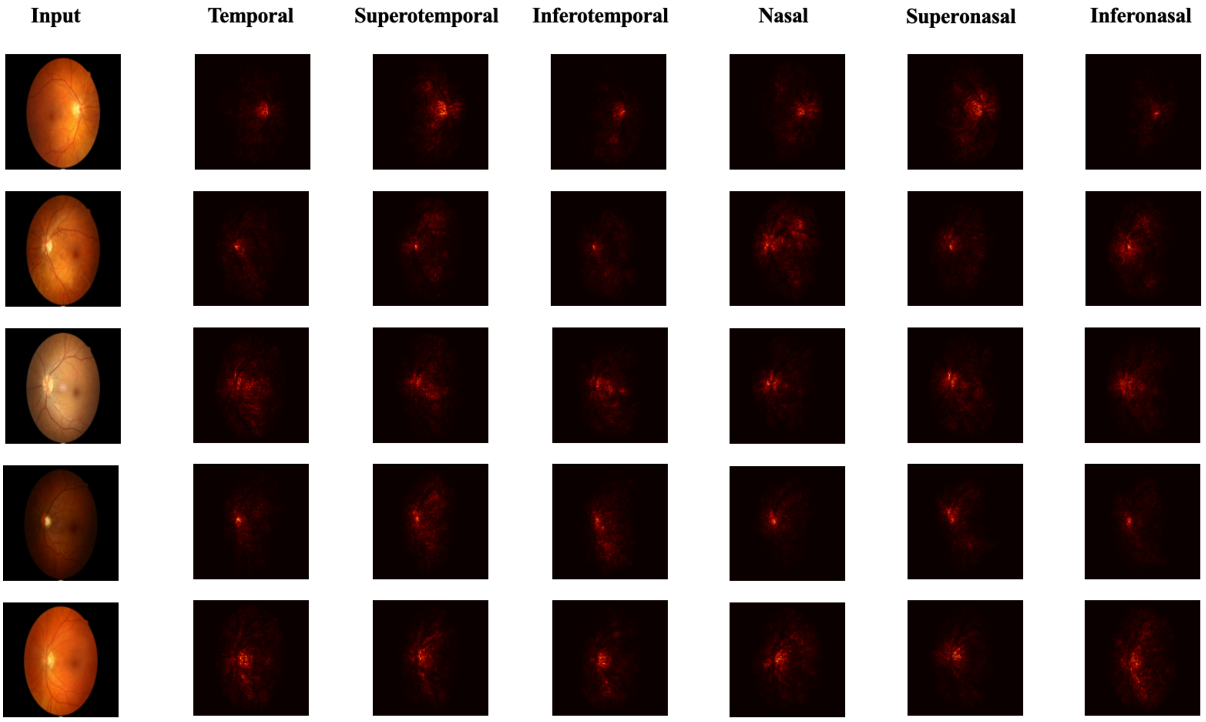
Input
Temporal
Superotemporal
Inferotemporal
Nasal
Superonasal
Inferonasal

**Supplementary Figure 3. Q-Q plots for AI-derived GWAS meta-analysis.** (a) Global pRNFL thickness, (b) Global BMO-MRW, (c) Sectoral pRNFL thickness, and (d) Sectoral BMO-MRW. Observed -log10(P-value) are plotted against expected -log10(P-value) under the null hypothesis of no association. The diagonal red line represents the null distribution. Inflation was assessed using the genomic inflation factor ($\lambda_{GC}$) and LD score regression (LDSC) intercept.

**(a) Global pRNFL thickness**

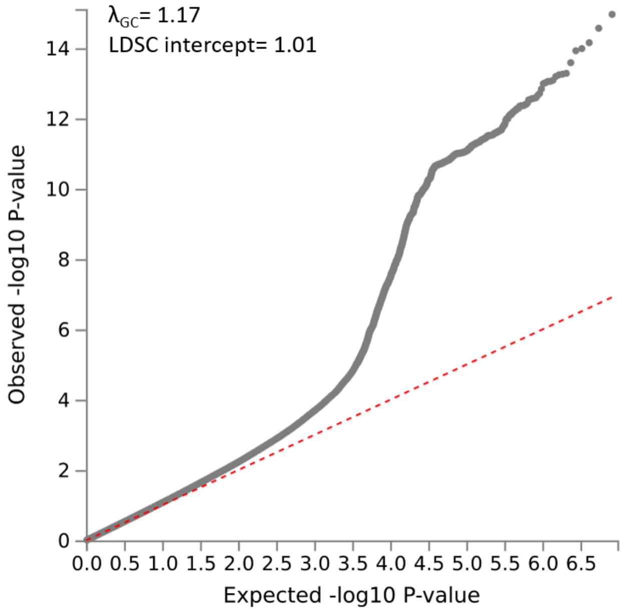


**(b) Global BMO-MRW**

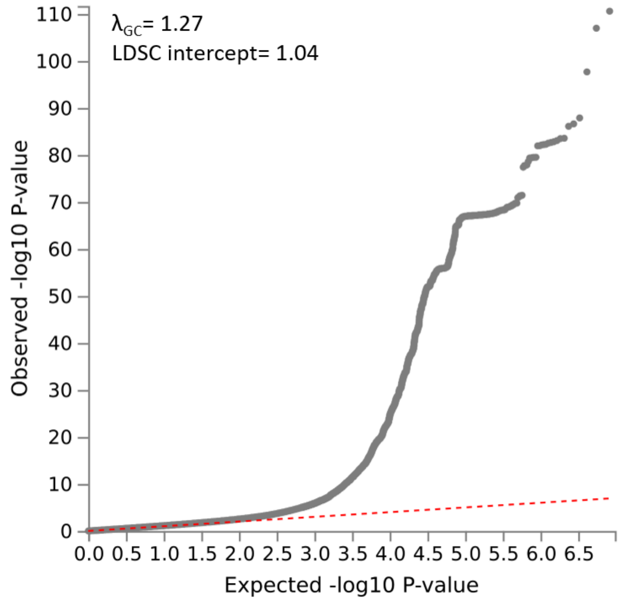

**(c) Sectoral pRNFL thickness**

Temporal

$\lambda_{GC}$ = 1.1747

LDSC intercept = 1.0271 (0.0077)

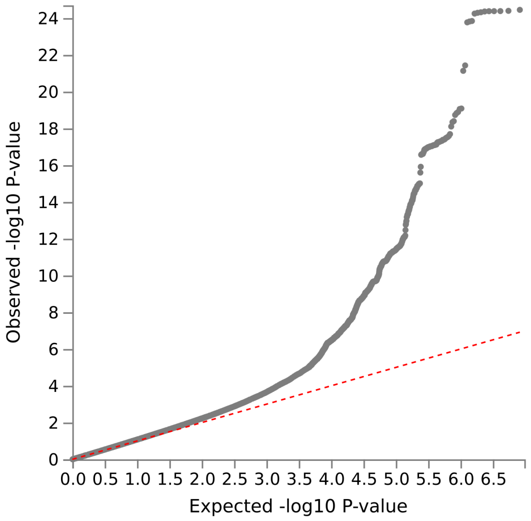


Nasal

$\lambda_{GC}$ = 1.127

LDSC intercept = 1.0094 (0.0078)

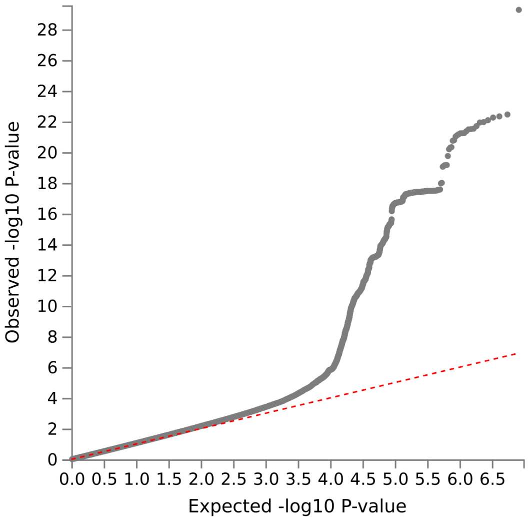


Superotemporal

$\lambda_{GC}$ = 1.1587

LDSC intercept = 1.0138 (0.0075)

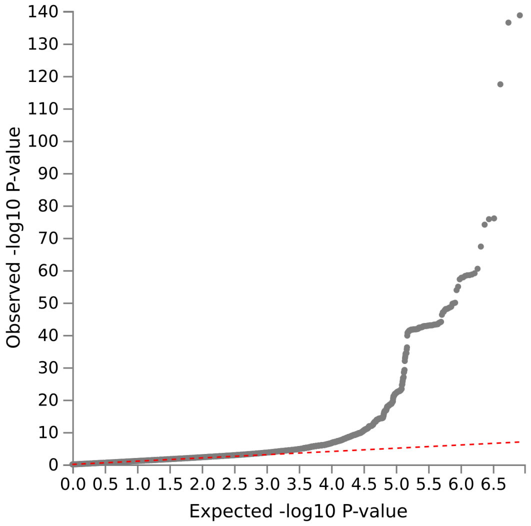


Superonasal

$\lambda_{GC}$ = 1.1651

LDSC intercept = 1.0052 (0.0079)

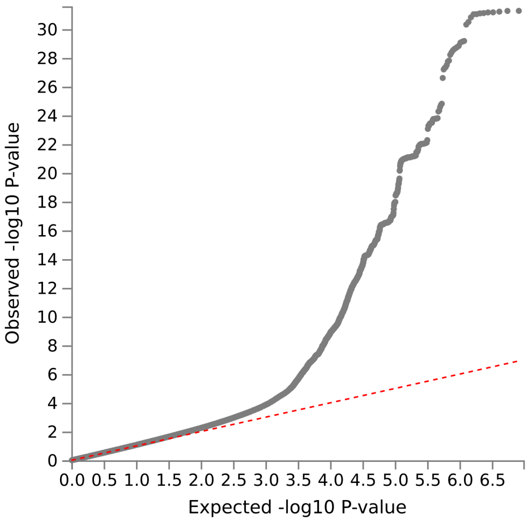


Inferotemporal

Inferonasal

$\lambda_{GC} = 1.1779$

LDSC intercept = 1.0202 (0.0077)

$\lambda_{GC} = 1.1715$

LDSC intercept = 1.0144 (0.0076)

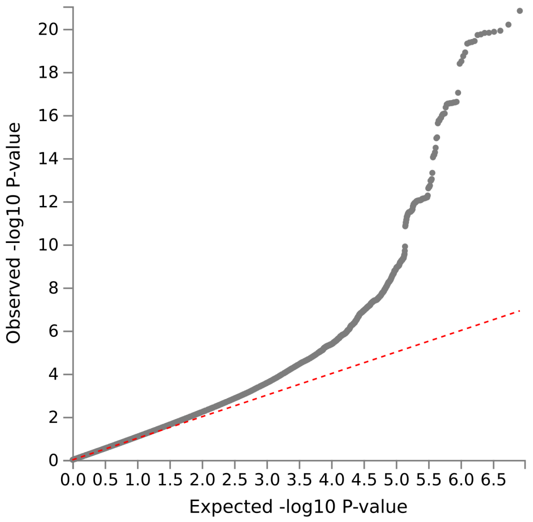


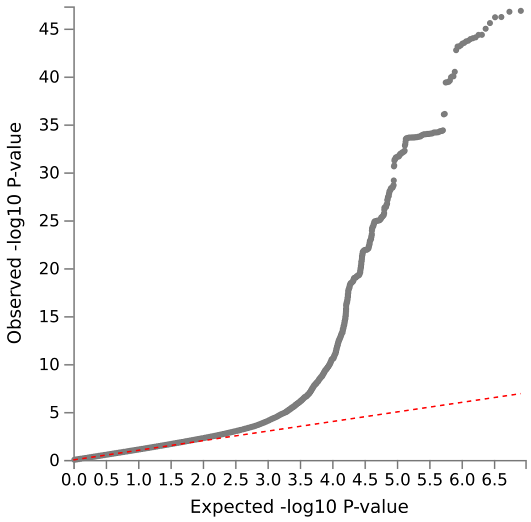


**(d) Sectoral BMO-MRW**

Temporal

Nasal

$\lambda_{GC} = 1.2365$

LDSC intercept = 1.0422 (0.0099)

$\lambda_{GC} = 1.2398$

LDSC intercept = 1.0413 (0.0098)

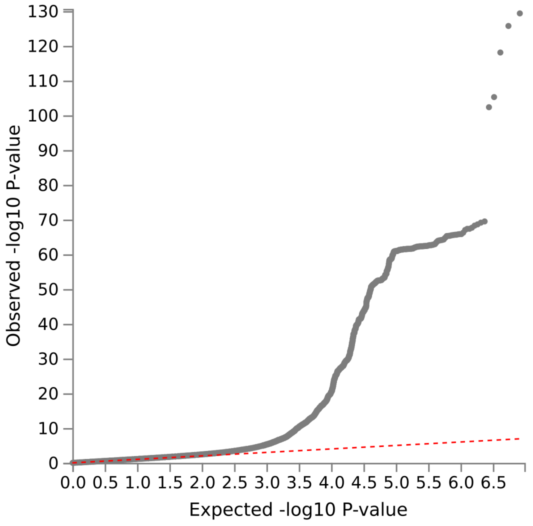


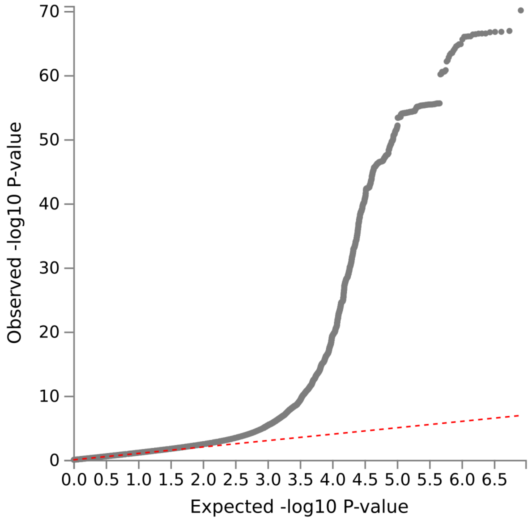


Superotemporal

Superonasal

$\lambda_{GC}$ = 1.2464

LDSC intercept = 1.0272 (0.0097)

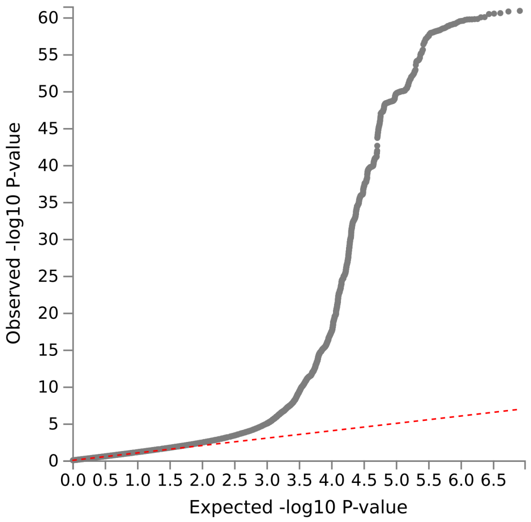


$\lambda_{GC}$ = 1.2498

LDSC intercept = 1.0413 (0.0099)

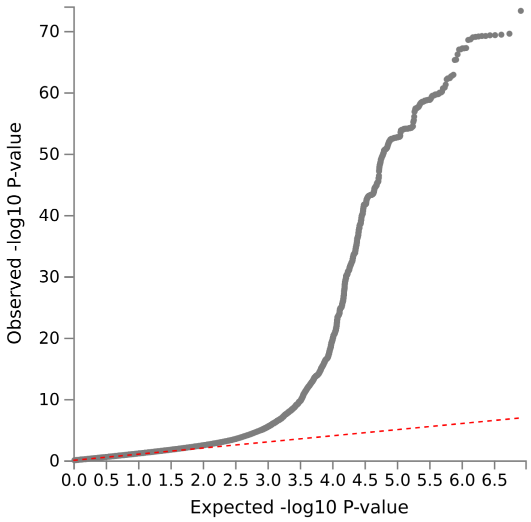


Inferotemporal

$\lambda_{GC}$ = 1.2299

LDSC intercept = 1.046 (0.0102)

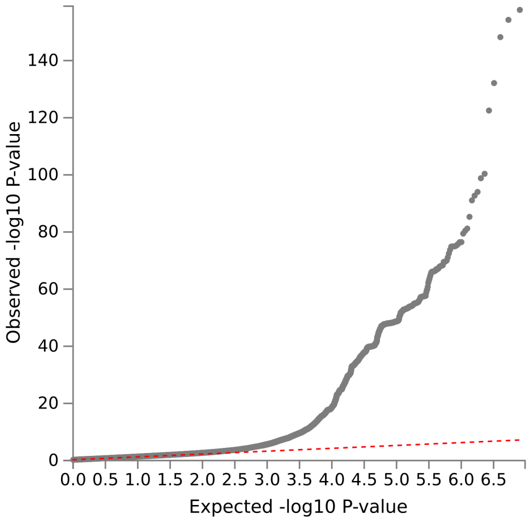


Inferonasal

$\lambda_{GC}$ = 1.2464

LDSC intercept = 1.041 (0.0103)

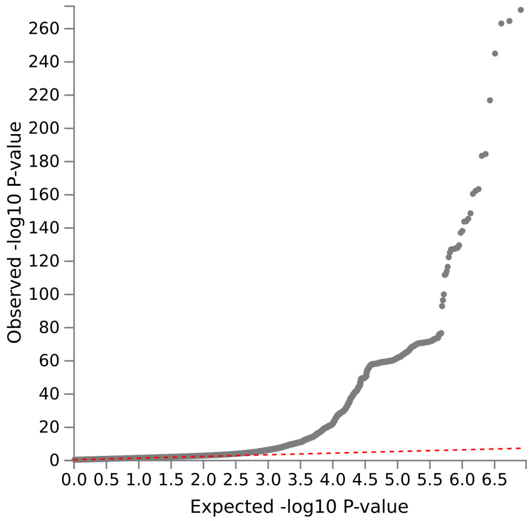

**Supplementary Figure 4. Manhattan plots for AI-derived GWAS meta-analysis.** (a) Sectoral pRNFL thickness and (b) Sectoral BMO-MRW. The -log10(P-value) of SNP associations are plotted against genomic position across chromosomes. The dashed red horizontal line represents the genome-wide significance threshold (P = 5 × $10^{-8}$). Each dot represents one SNP. Genome-wide significant loci were defined using LD clumping ($r^2 < 0.2$ within 1 Mb).

**(a) Sectoral pRNFL thickness**

Temporal

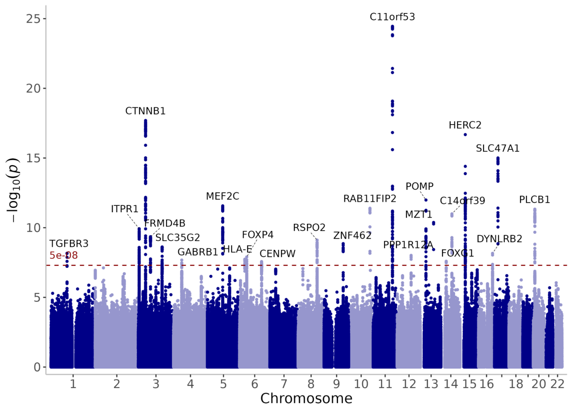


Nasal

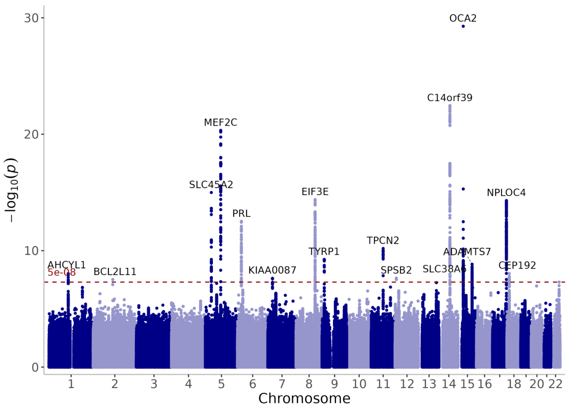


Superotemporal

Superonasal

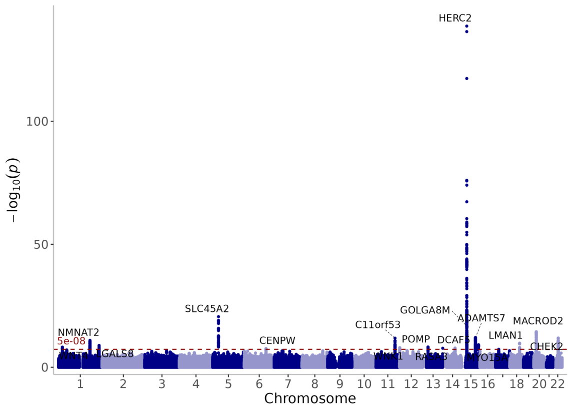


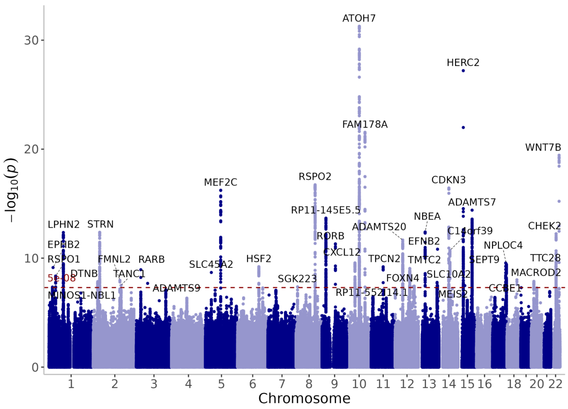


Inferotemporal

Inferonasal

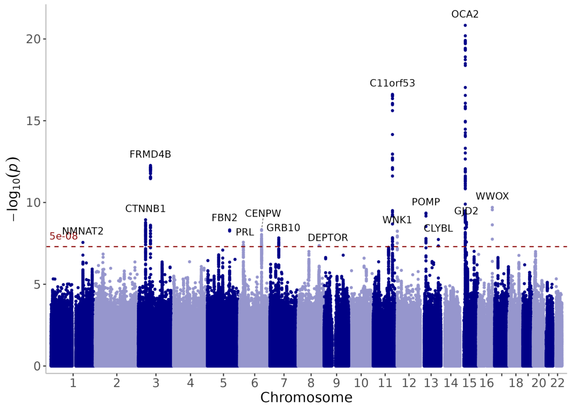


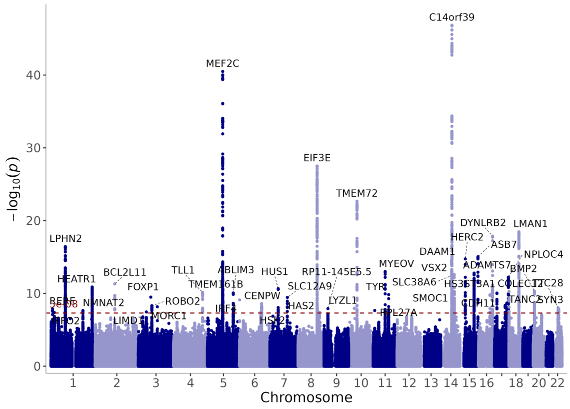


### (b) Sectoral BMO-MRW

Temporal

Nasal

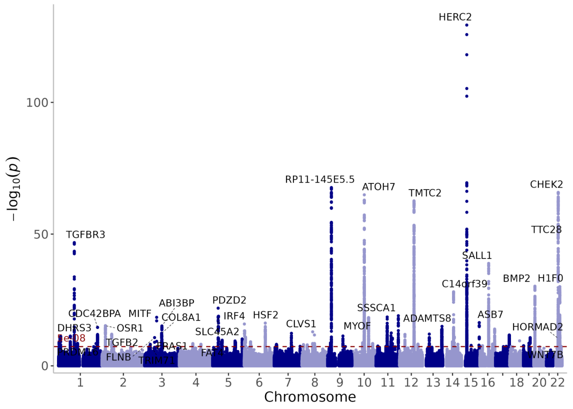


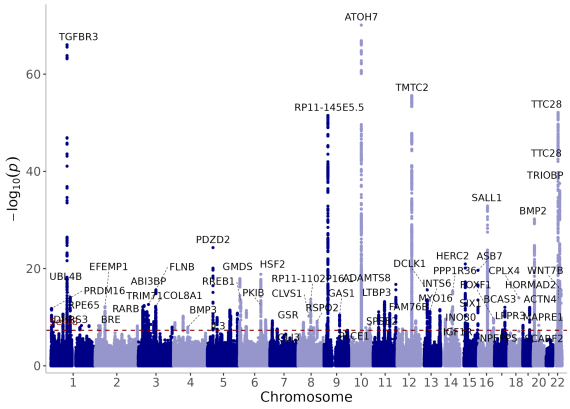


Superotemporal

Superonasal

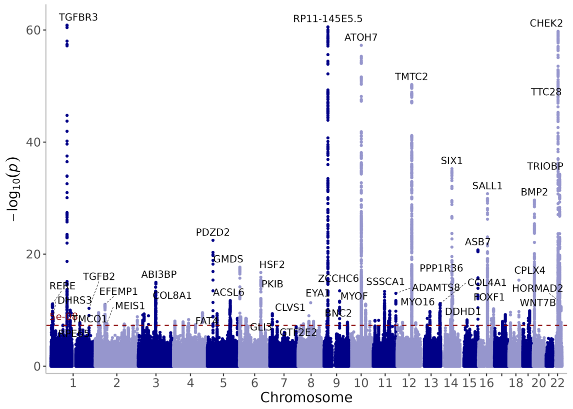


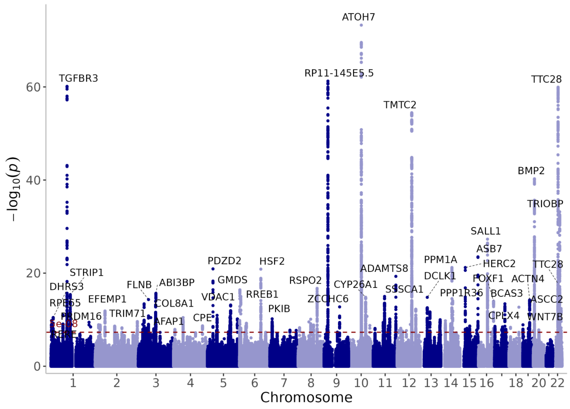


Inferotemporal

Inferonasal

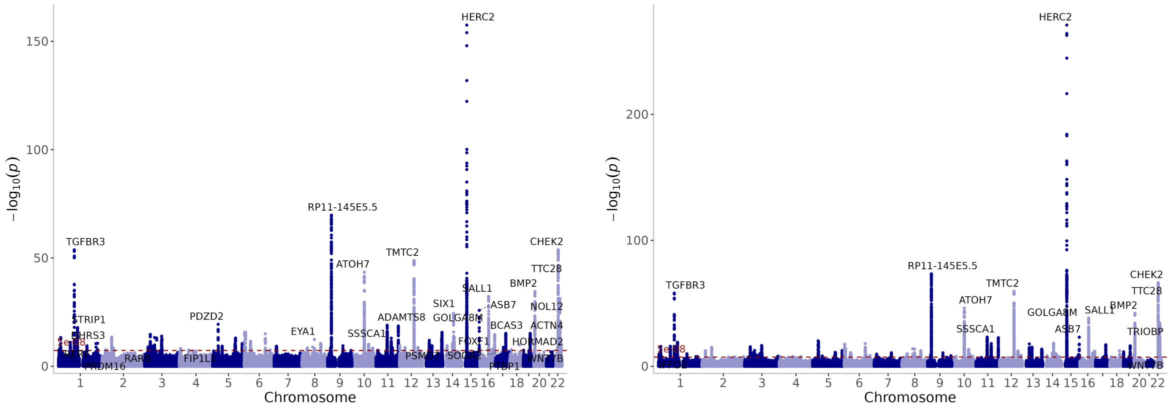

−log10(p)
Chromosome
HERC2
RP11-145E5.5
TGFBR3
CHEK2
TMTC2
ATOH7
TTC28
BMP2
SALL1
ASB7
NOL12
SIX1
GOLGA8M
ADAMTS8
PDZD2
STRIP1
BCAS3
ACTN4
EYA1
SSSCA1
FOXF1
HORMAD2
RARB
FIP1L1
PRDM16
WNT7B
TRIOBP
1e-08

**Supplementary Figure 5. Concordance of SNP effect sizes between AI-derived and directly measured thickness GWAS.** (a) Sectoral pRNFL thickness and (b) Sectoral BMO-MRW. Scatter plots compare effect size estimates (β) for lead SNPs at genome-wide significant loci ($P < 5 \times 10^{-8}$) identified in the AI-derived GWAS (x-axis) and in GWAS of directly measured thickness in IGGC cohorts (y-axis). Horizontal and vertical error bars around each lead SNP represent 95% confidence intervals for effect size estimates in the AI-derived and IGGC GWAS, respectively. The blue line indicates the linear regression fit. Each point represents one lead SNP. Pearson's correlation coefficient (r) with the corresponding P value is shown.

**(a) Sectoral pRNFL thickness**

Temporal

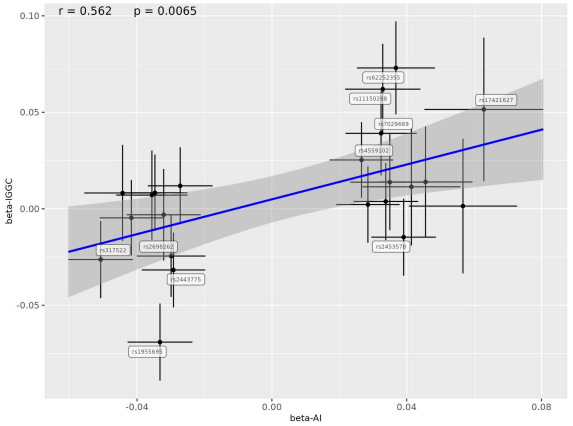


Nasal

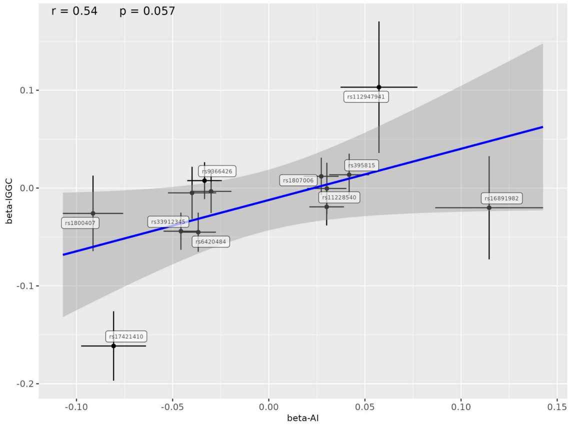


Superotemporal

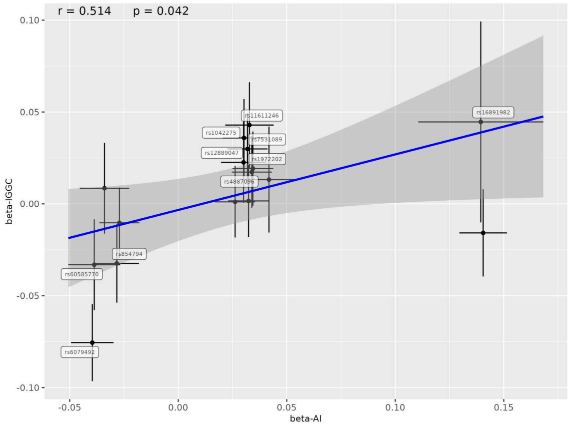


Superonasal

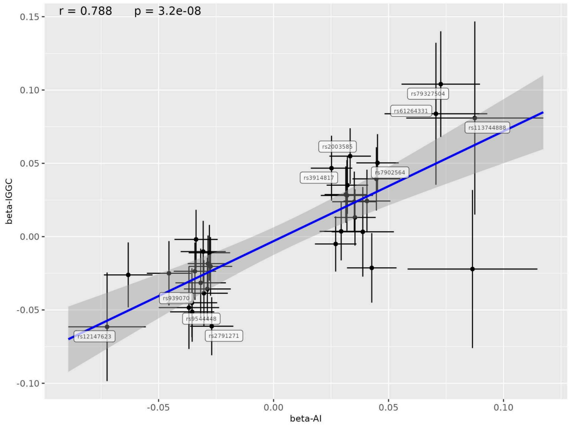


Inferotemporal

Inferonasal

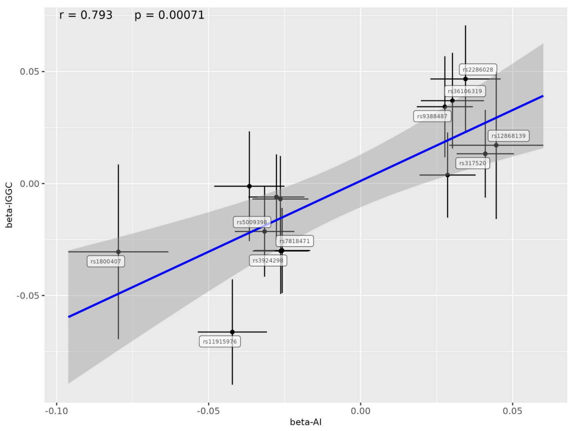


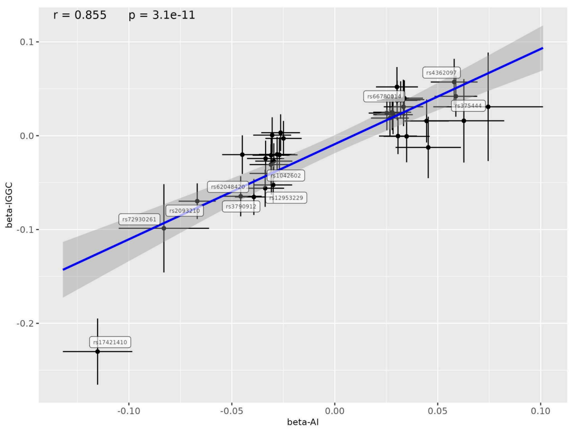


#### (b) Sectoral BMO-MRW

Temporal

Nasal

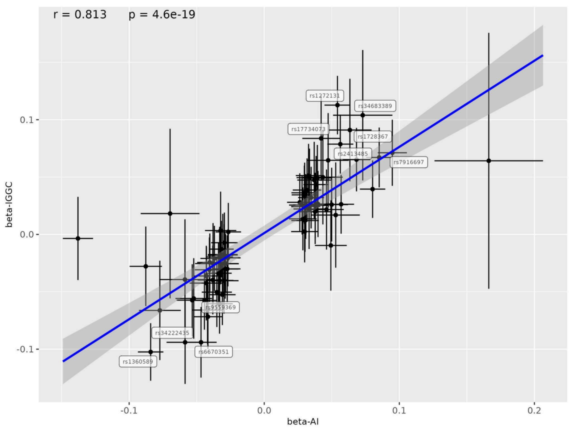


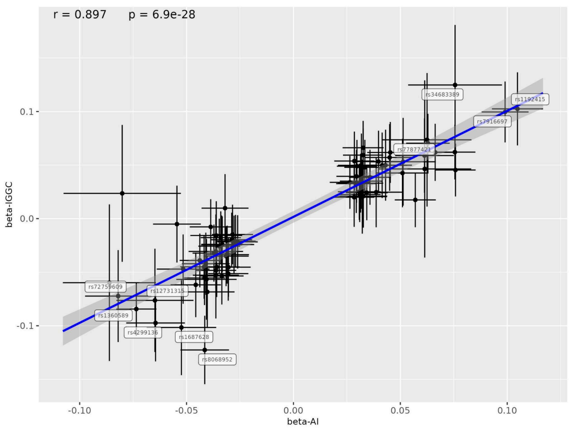


Superotemporal

Superonasal

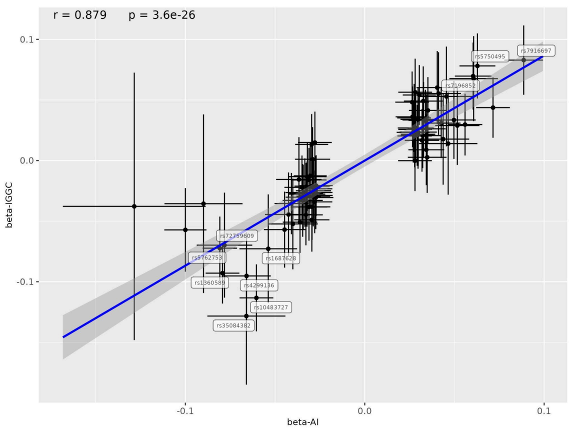


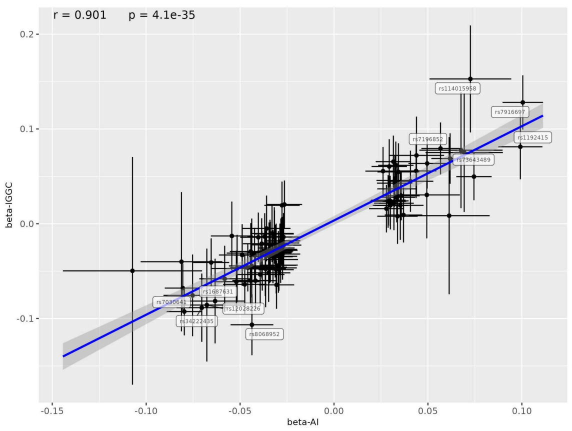


Inferotemporal

Inferonasal

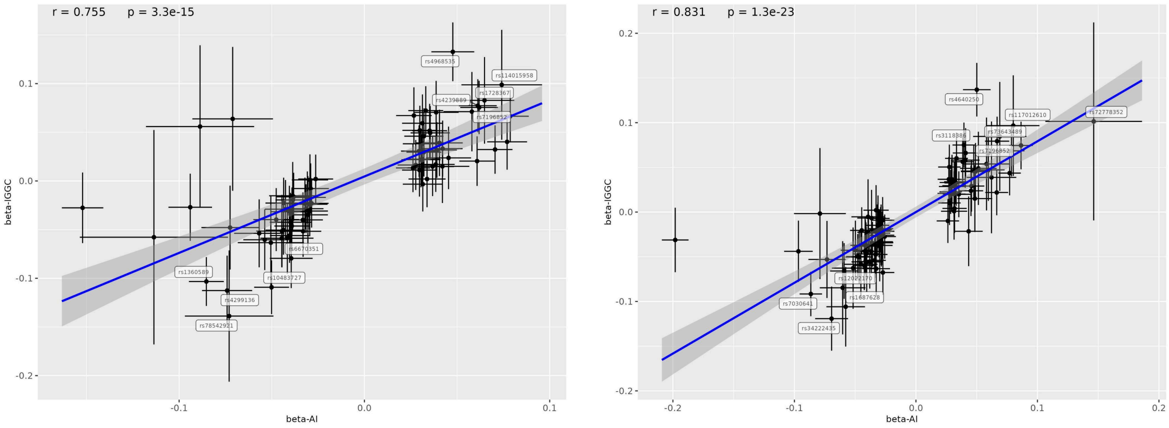

r = 0.755 p = 3.3e-15
beta-IGGC
beta-AI
rs4968535
rs114015958
rs1728367
rs4239889
rs7196852
rs6670351
rs1360589
rs10483727
rs4299136
rs78542921
r = 0.831 p = 1.3e-23
beta-IGGC
beta-AI
rs4640250
rs117012610
rs72778352
rs3118386
rs73643489
rs7196852
rs12022170
rs1687628
rs7030641
rs34222435

**Supplementary Figure 6. Manhattan plots for AI-derived and directly measured thickness MTAG analysis.** (a) Sectoral pRNFL thickness and (b) Sectoral BMO-MRW. The -log10(P-value) of SNP associations are plotted against genomic position across chromosomes. The dashed red horizontal line represents the genome-wide significance threshold ($P = 5 \times 10^{-8}$). Each dot represents one SNP. Genome-wide significant loci were defined using LD clumping ($r^2 < 0.2$ within 1 Mb).

**(a) Sectoral pRNFL thickness**

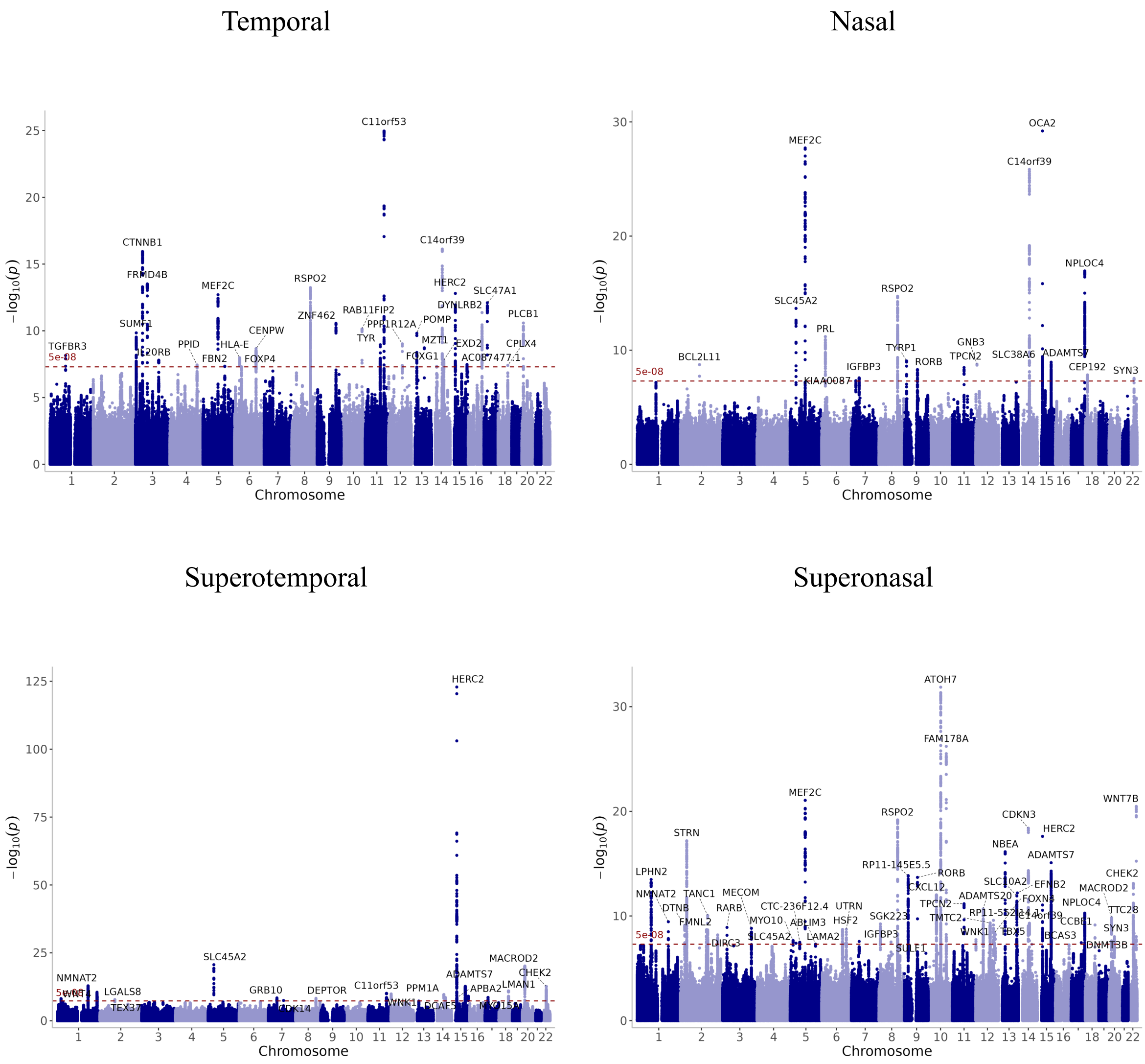


Inferotemporal

Inferonasal

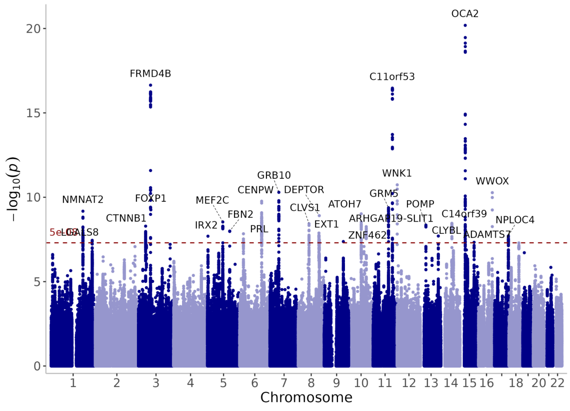


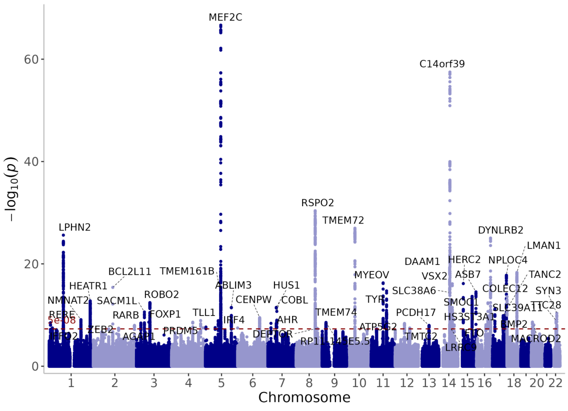


#### (b) Sectoral BMO-MRW

Temporal

Nasal

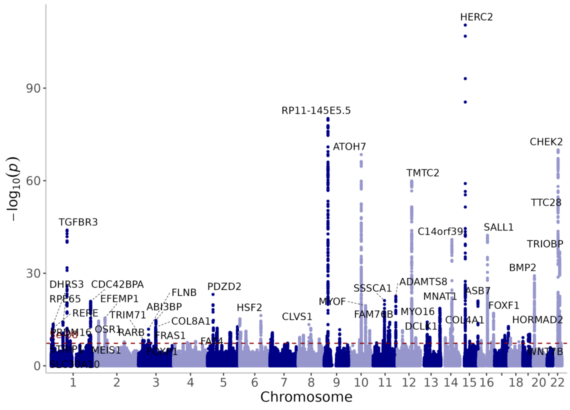


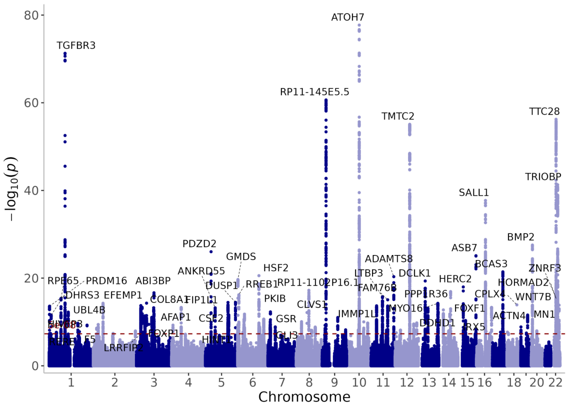


Superotemporal

Superonasal

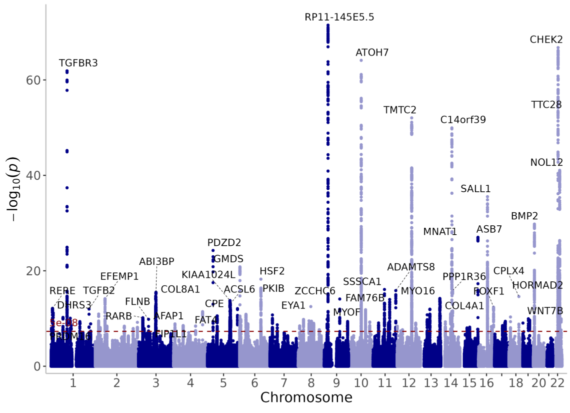


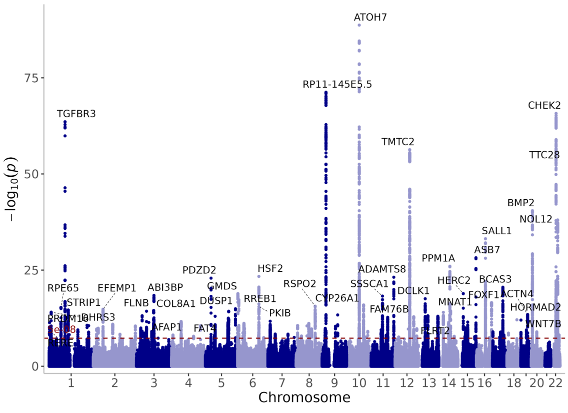

Inferotemporal    Inferonasal

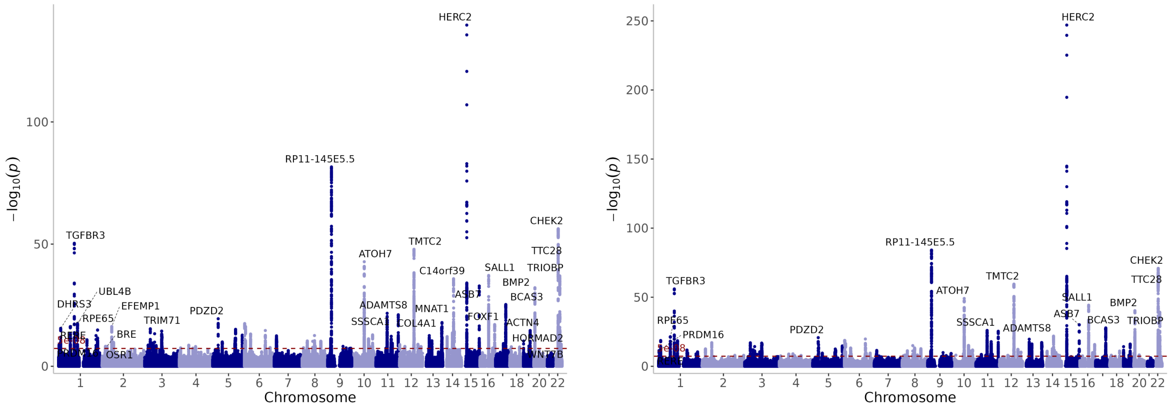
